# Metastability as a neuromechanistic biomarker of schizophrenia pathology

**DOI:** 10.1101/2022.10.14.22281093

**Authors:** Fran Hancock, Fernando E. Rosas, Robert A. McCutcheon, Joana Cabral, Ottavia Dipasquale, Federico E. Turkheimer

## Abstract

The disconnection hypothesis of schizophrenia proposes that symptoms of the disorder arise as a result of aberrant functional integration between segregated areas of the brain. The concept of metastability characterizes the coexistence of competing tendencies for functional integration and functional segregation in the brain and is therefore well suited for the study of schizophrenia. In this study we investigate metastability as a neuromechanistic biomarker of schizophrenia pathology, including a demonstration of reliability and face validity.

Group-level discrimination, individual-level classification, and pathophysiological relevance were assessed using two independent case-control studies of schizophrenia, the Human Connectome Project Early Psychosis (HCPEP) study (controls *n*=82, non-affective psychosis *n*=53) and the Cobre study (controls *n*=71, cases *n*=59). In this work we introduce a new framework that uses Leading Eigenvector Dynamic Analysis (LEiDA) to capture specific features of dynamic functional connectivity and then implements a novel approach to estimate metastability. We used non-parametric testing to evaluate group-level differences and a naïve Bayes classifier to discriminate cases from controls.

Our results show that our new approach is capable of discriminating cases from controls with elevated effect sizes relative to published literature, reflected in an up to 76% area under the curve (AUC) in out-of-sample classification analyses. Furthermore, our analyses demonstrated that patients with early psychosis exhibit intermittent disconnectivity of subcortical regions with frontal cortex and cerebellar regions, introducing new insights about the mechanistic bases of these conditions.

Overall, these findings demonstrate reliability and face validity of metastability as a neuromechanistic biomarker of schizophrenia pathology.

## Introduction

Schizophrenia affects roughly 1% of the population, is associated with premature mortality and morbidity, and is accompanied by a large social and financial burden [1]. While schizophrenia can be a chronic disorder for a significant proportion of individuals [2], there is evidence that early diagnosis and treatment can lead to improved outcomes for patients [3]. Biomarkers of schizophrenia in early and established phases may differ, and hence may be informative of developing pathophysiology.

The disconnection hypothesis of schizophrenia states that the disorder can be understood as a failure of functional integration in the brain. Functional integration is closely related with the functional connectivity, and with the influence of brain dynamics of one region on another [4,5]. Failure of functional integration manifests as a disruption of the coordination required for the normal functioning of distributed brain regions [6]. For example, abnormal functioning of the basal ganglia in schizophrenia has previously been found with fMRI in schizophrenia [7–10]. Indeed, ganglia hyperdopaminergia may be attributable to disconnectivity stemming from GABA parvalbumin interneuron disorder [11].

Disconnection in schizophrenia has been investigated with static functional connectivity (FC) [12–15]. However, static FC relies on statistical relationships between fMRI signals throughout the complete scan, which forces it to discard critical information about the brain’s dynamics. In contrast, it is reasonable to believe that dynamic approaches – which consider the temporal dynamics of fMRI signals - may have the potential to discover more precise and informative biomarkers [16–29]. Unfortunately, the literature provides no empirical studies investigating if approaches which rely on collective dynamical properties have better classification ability than those that rely on static FC properties, and whether dynamical approaches provide relevant insight for biological and cognitive interpretation.

To address this important issue, in this work we analyze the suitability of a specific marker of brain dynamics: *metastability*. Metastability is a concept originating from dynamical systems theory which provides an explanation for the spontaneous and self-organized emergence and dissolution of spatiotemporal patterns of coordination activity [30,31]. In a neuroscientific context this reflects a tension established by the competition between trends for functional specialization and functional integration within and between brain regions [32]. Metastability is nowadays an ubiquitous concept across diverse models of brain functioning including coordination dynamics [33] and complex systems [34], while its metrics have found application in both empirical studies and computational modeling [35–44]. Furthermore, a proxy measure of metastability was recently found to be stable and representative across multiple fMRI scans of healthy young adults, highlighting its potential as a group-level biomarker of psychiatric disorders [42].

Building on this previous work, here we investigate how metastability would perform as a neuromechanistic biomarker of schizophrenia at the group- and individual-level; if this performance would carry over to face validation; and what this putative biomarker would tell us about the pathophysiology of schizophrenia.

We introduce a new measure for metastability as the mean variance of instantaneous phase-locking. Our rationale for this operationalization stems from the theory of Synergetics [45] and recent generalization of the Haken-Kelso-Bunz (HKB) model to multiple oscillators [46], which exhibits stable antiphase synchronization [47], and from the observation that differences in connectivity were not reflected in differences in the traditional measure for metastability within this study. We found that this novel proxy for metastability distinguished patients with established schizophrenia from healthy controls at the group-level with moderate effect size (*d* = 0.77), delivered performance in the range of published individual-level classifiers for cross-validation, and out-of-sample testing, and highlighted dysfunctional connectivity in basal ganglia in early schizophrenia, and so demonstrated face validity of metastability as a neuromechanistic biomarker schizophrenia pathology.

## Results

### Derivation of spatiotemporal patterns of phase-locking

We analyzed the resting-state fMRI activity from a total of 670 scanning sessions from the Human Connectome Project Early Psychosis (HCPEP) and Cobre datasets (see Materials and methods). In the HCPEP dataset healthy controls (CON, *n*=53) and subjects with non-affective psychosis (NAP, *n*=82) participated in 4 scanning sessions on 2 consecutive days. In the Cobre dataset CON (*n*=71) and subjects with schizophrenia (SCHZ, *n*=59) participated in 1 scanning session. Each dataset consisted of whole-brain fMRI signals averaged over *n*=116 cortical, subcortical, and cerebellar brain regions as defined in the AAL116 anatomical parcellation [48].

We used instantaneous phase-locking to measure the interaction between fMRI signals related to different brain regions. The fMRI time-series of each subject was filtered within the narrowband 0.01-0.08 Hz which did not violate the Bedrosian Theorem (see Materials and methods) [42]. The filtered signal was then transformed into amplitude and phase via the Hilbert transform, and the resulting phase time series was analyzed via the Leading Eigenvector Dynamic Analysis (LEiDA) [42]. In order to identify recurrent spatiotemporal patterns of phase-locking – henceforth called ‘LEiDA modes’ – we performed k-means clustering on the phase-locked time-series of each of the datasets that were analyzed (HCPEP CONx4, HCPEP NAPx4, Cobre CONx1, Cobre SCHZx1, see Materials and methods). We calculated the results for *k*=2-10 clusters, and then choose *k*=5 LEiDA modes - denoted as *ψ*_1_, *ψ*_2_, *ψ*_3_, *ψ*_4_, *ψ*_5_-according to silhouette values [49] (see S1 Fig), which is consistent with previous studies [42,50,51]. Additionally, we calculated the instantaneous magnetization as the ratio of in-phase regions to anti-phase regions, which indicates criticality [52]. Fig 1 shows the diversity of phase-locking behavior for two individual subjects from the HCPEP dataset.

**Fig 1.**
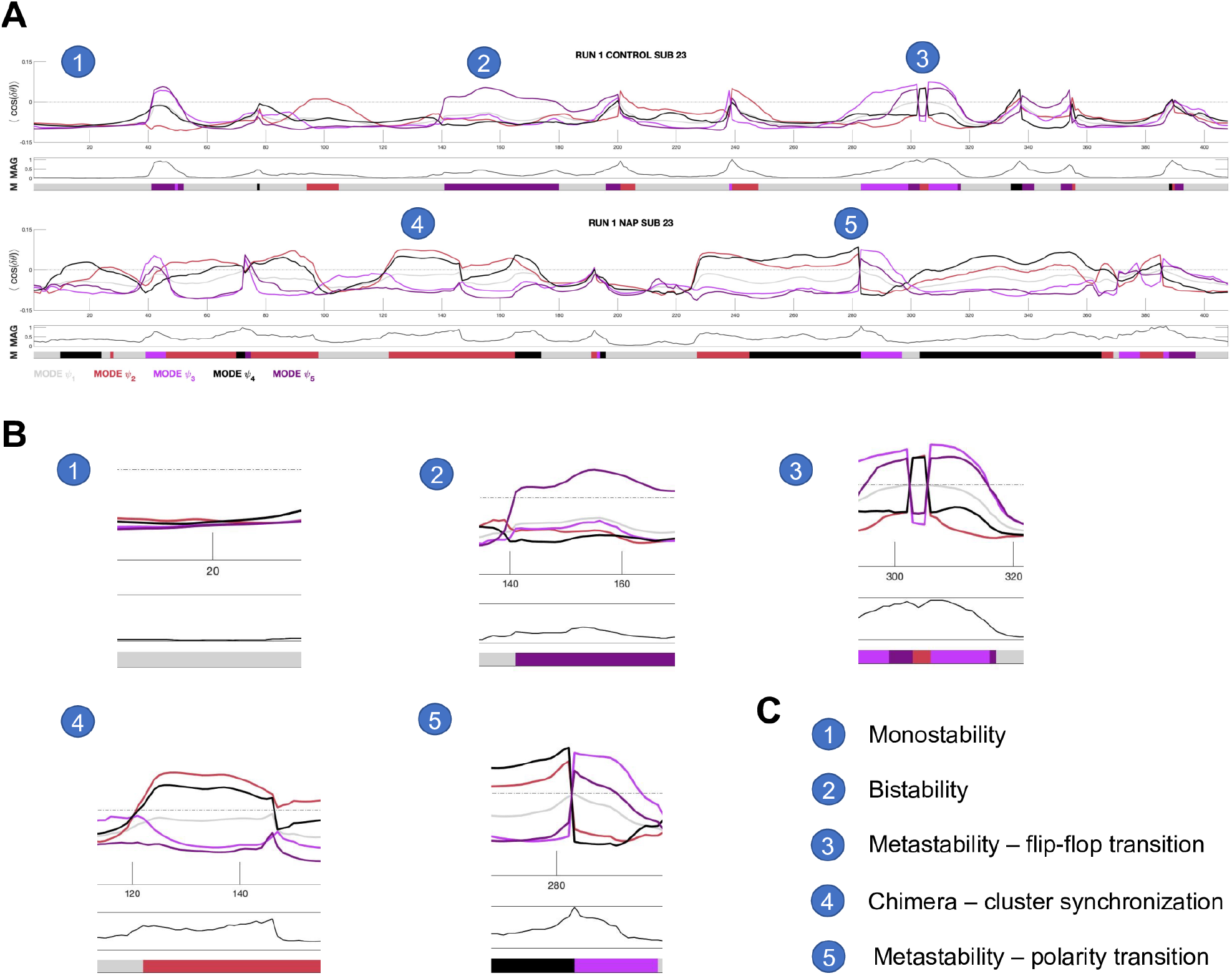
Diversity of phase-locking behavior. **A)** Timeseries of mode eigenvectors from two subjects from the HCPEP dataset. Top panel shows phase-locking behavior. Middle panel shows instantaneous magnetization which is the ratio of in-phase to anti-phase regions. Bottom panel shows the mode assigned to the timepoint from k-means clustering. Interesting behavior is indicated with numbered circles. **B)** Blow-outs for points 1 to 5. **C)** Legend for the numbered circles. MAG, magnetization; M, mode. Gray dotted line shows where phase-locking is equal to zero.

We found that the 5 modes reflected connectivity within and across known resting-state networks, subcortical and cerebellar regions. Following Ref. [42], we visualized each mode in 10mm voxel space by averaging the eigenvector values over all time instances assigned to a particular mode. We visualized FC as connectograms by taking the FC matrices for each mode and retaining regions that were collectively in-phase but out-of-phase with the global mode (see Fig 2)

**Fig 2.**
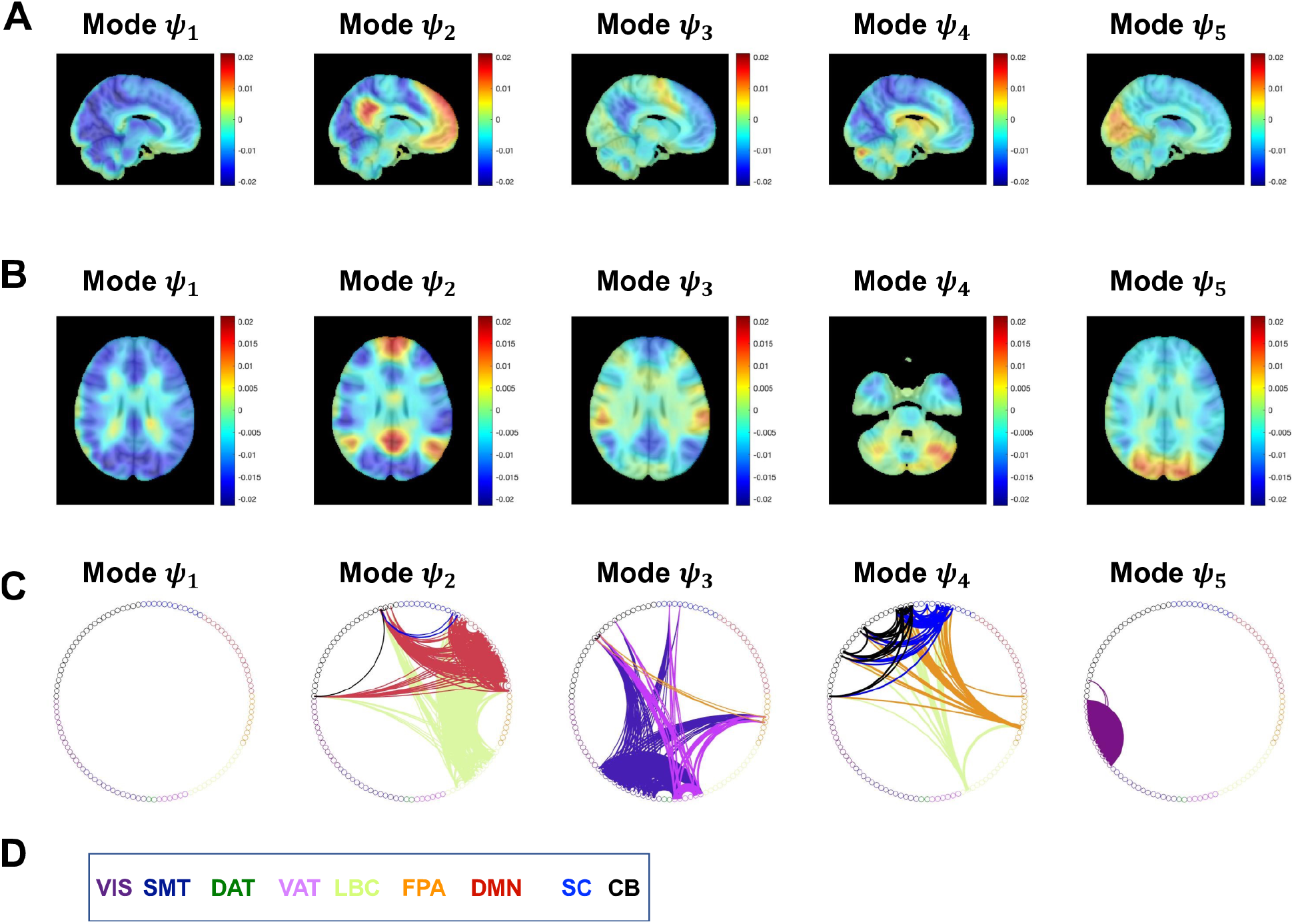
Spatial patterns of recurrent phase-locked connectivity in run 3 for controls. **A)** Phase-locking patterns for the 5 modes in sagittal view. **B)** Phase-locking patterns for the 5 modes in axial view. **C)** Respective FC presented as connectograms color-coded as in Yeo [53] with the addition of dark blue for subcortical regions, and black for cerebellar regions. In Mode *ψ*_1_ all regions are aligned in-phase and so there is no anti-phase connectivity. FC computed as the outer product of the leading eigenvector for each mode. **D)** Color coded legend for the Yeo resting-state networks, subcortical and cerebellar regions. VIS, Visual; SMT, Somatomotor; DAT, Dorsal attention; VAT, Ventral attention; LBC, Limbic; FPA, Frontal parietal; DMN, Default mode network; SC, Subcortical; CB, Cerebellar.

Using the modes from RUN 3 in CON as an illustrative example, we find that Mode *ψ*_1_ represents a global mode where the fMRI signals in all regions are aligned in-phase without anti-phase connectivity. Mode *ψ*_2_ exhibits connectivity within Default Mode Network (DMN), Limbic network (LBC), and cerebellum (CB), and connectivity between DMN-LBC, DMN-subcortical (SC), DMN-CB, LBC-SC, LBC-CB. Mode *ψ*_3_ shows connectivity within Somatomotor (SMT), Ventral Attention network (VAT), Frontal Parietal Area (FPA) and CB, and connectivity between SMT-FPA, SMT-CB, SMT-CB, VAT-FPA, VAT-SC, VAT-CB and FPA-CB. Mode *ψ*_4_ exhibits connectivity within SC and CB, and connectivity between LCB-FPA, LBC-SC, LBC-CB, FPA-SC, FPA-CB, and SC-CB. Finally, Mode *ψ*_5_ shows connectivity within Visual network (VIS), and between VIS-CB.

### Characteristics of spatiotemporal modes

Before assessing differences in the modes across the case-control groups, we first controlled if the modes observed in HCPEP were stable and representative across the four runs. We calculated run reliability within groups with interclass correlation ICC(1,1) [54] (See Materials and methods). The modes extracted for CON showed substantial to almost perfect reliability between runs with median ICC values *ψ*_1_ (0.96), *ψ*_2_ (0.98), *ψ*_3_ (0.64), *ψ*_4_ (0.89), and *ψ*_5_(0.77) The modes extracted for NAP also showed substantial to almost perfect reliability with median ICC values *ψ*_1_ (0.97), *ψ*_2_ (0.97), *ψ*_3_ (0.96), *ψ*_4_ (0.77), and *ψ*_5_(0.82) (see S2 Fig for all ICC matrices). We therefore confirmed that the modes tended to be invariant across multiple acquisitions in both case and control groups in HCPEP.

Concentrating first on HCPEP, we found that there was a strong contribution of basal ganglia regions to the leading eigenvector for Mode *ψ*_4_ in CON. We therefore assessed if there were differences in basal ganglia connectivity, measured as contribution to Mode *ψ*_4_, between the groups. Regional contribution was calculated as the mean value of instantaneous phase-locking over time for the region of interest (ROI). We first investigated group (CON, NAP), run (RUN1, RUN2, RUN3, RUN4), and interactions between group and run on bilateral caudate, putamen, pallidum, and thalamus. Using a 2×4 non-parametric ANOVA with the Aligned Rank Transform (ART) [55,56], we found significant interactions between group and run (Table 1).

**Table 1.**
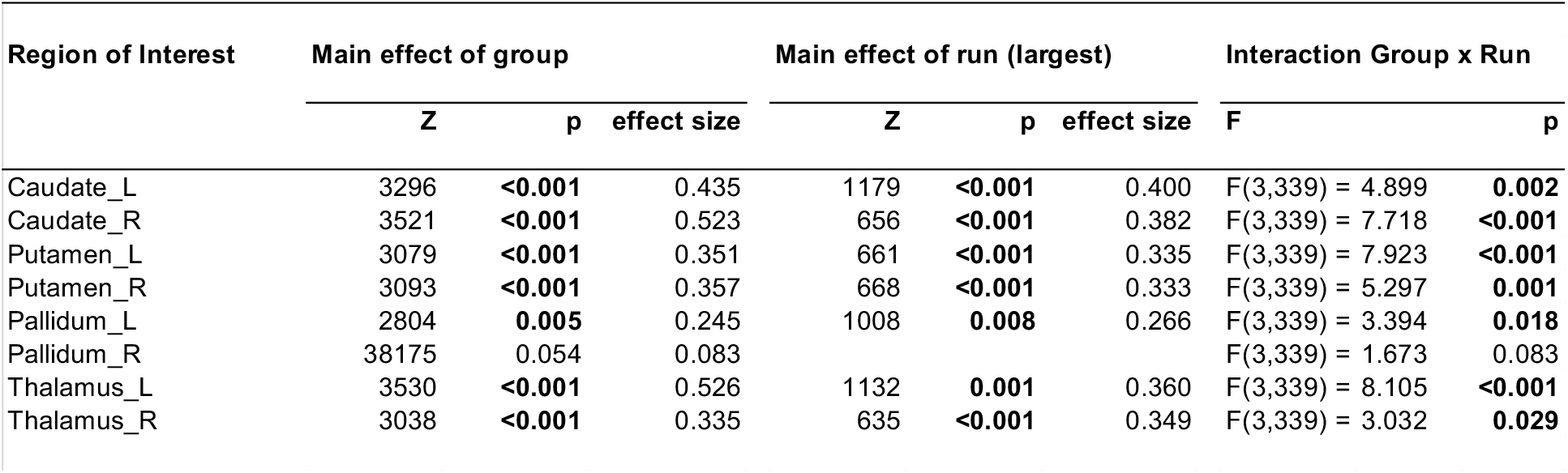
Effects of group, run, and interactions between group and run, on contributions to mode *ψ*_4_ connectivity in the bilateral caudate, putamen, pallidum, and thalamus. Bold font indicates statistical significance following Bonferroni correction for multiple comparisons.

We found significant main effects of run in both groups for multiple ROIs. The effects and the drivers of these effects are detailed in S1 Supporting Information. The largest main effects of run are shown in Table 1.

Furthermore, we found significant main effects of group in Caudate_L, Caudate_R, Putamen_L, Putamen_R, Pallidum_L, Thalamus_L, and Thalamus_R (Table 1). We retained only group differences that were greater than these run effects. We thus found significant group differences in RUN2 for Caudate_L (*p*<0.001, *effect size*= 0.435), Caudate_R (*p*<0, *effect size*= 0.523), Putamen_L (*p*<0.001, *effect size*=0.351), Putamen_R (*p*<0.001, *effect size*=0.357) and Thalamus_L (*p*<0.001, *effect size*=0.526).

We therefore inferred that these group differences in basal ganglia contribution in RUN2 are not due to run effects, and indeed reflect group differences in regional contribution to Mode *ψ*_4_. (See Fig 3 and S1 Supporting Information for complete results of the statistical testing).

**Fig. 3.**
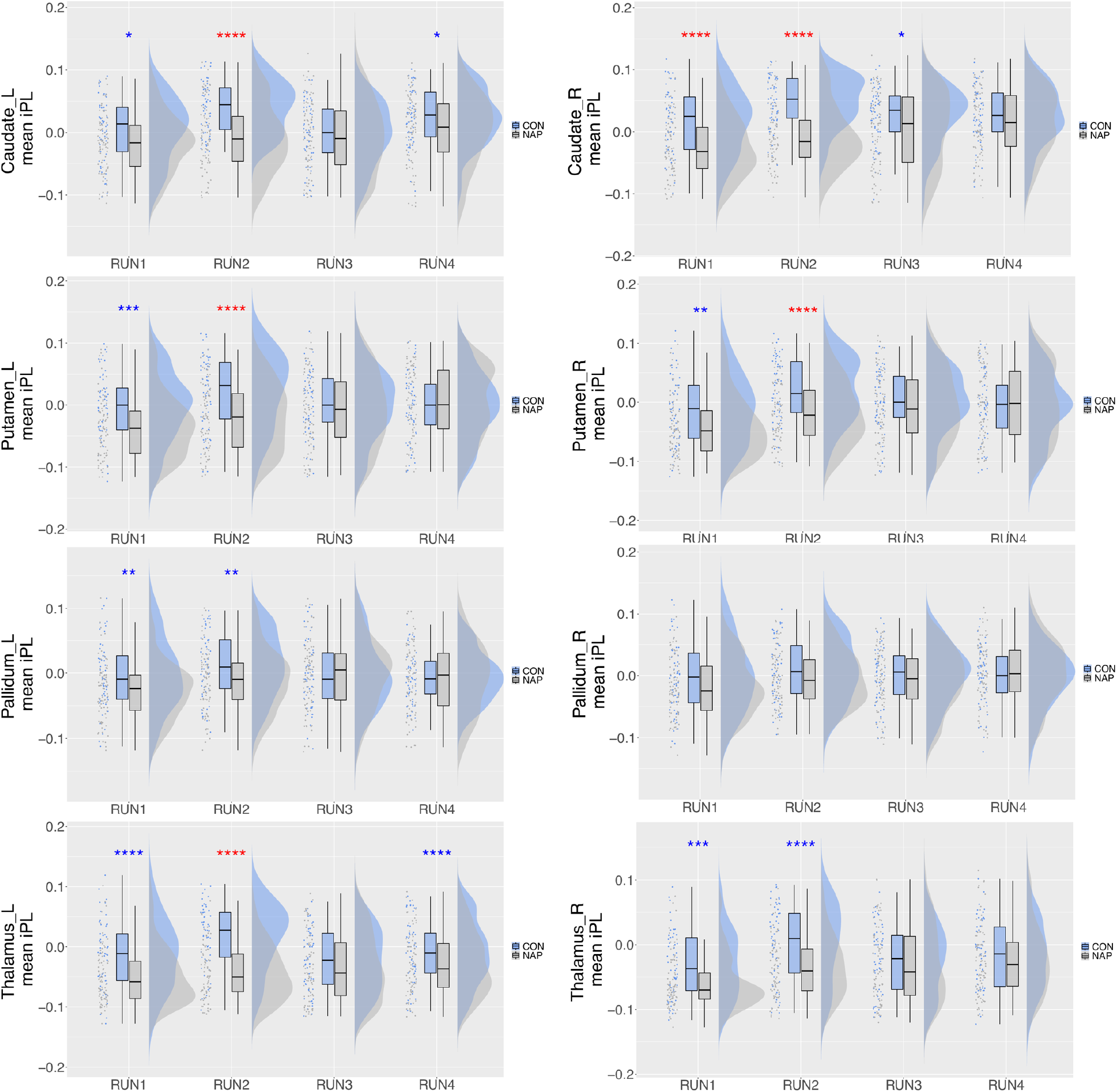
Group differences in regional contribution to the leading eigenvector for Mode *ψ*_4_. Regional contribution was calculated as the mean value of instantaneous phase-locking over time for a particular anatomical region of interest. Raincloud plots show from left to right scatter plot for the raw data, boxplots showing the median, upper and lower quartiles, upper and lower extremes, and the distributions of the raw data. iPL, instantaneous phase-locking, *=0.05, **=0.01, ***=0.001, ****<0.001. Red * effect size between groups greater than effect size between runs. Blue * effect size between groups less than largest effect size between runs.

### Global and local metastability – group-level neuromechanistic biomarkers of schizophrenia

To assess the performance of metastability at group-level, we computed and analyzed differences within and between groups based on the standard estimators for global and local metastability [57] (see S1 Supporting Text for the analysis, and S2 Supplementary Information for complete statistical results). We were somewhat surprised that metastability in Mode *ψ*_4_ was not significantly different between groups in HCPEP given the differences found in basal ganglia connectivity. On reflection, we realized that the modes were extracted based on phase-locking, whilst metastability was computed on phase synchrony. In other words, the standard deviation of phase synchrony only captured the variability of the in-phase regions and ignored the anti-phase regions. To rectify this methodological difference, we defined a new proxy for metastability as the mean variance of instantaneous phase-locking, VAR (See Materials and methods).

For the HCPEP dataset we first investigated group (CON, NAP), run (RUN1, RUN2, RUN3, RUN4), and interactions between group and run, on global VAR. Using a 2×4 non-parametric ANOVA with the Aligned Rank Transform (ART) [55,56], we found a significant interaction between group and run (Table 2).

**Table 2.**
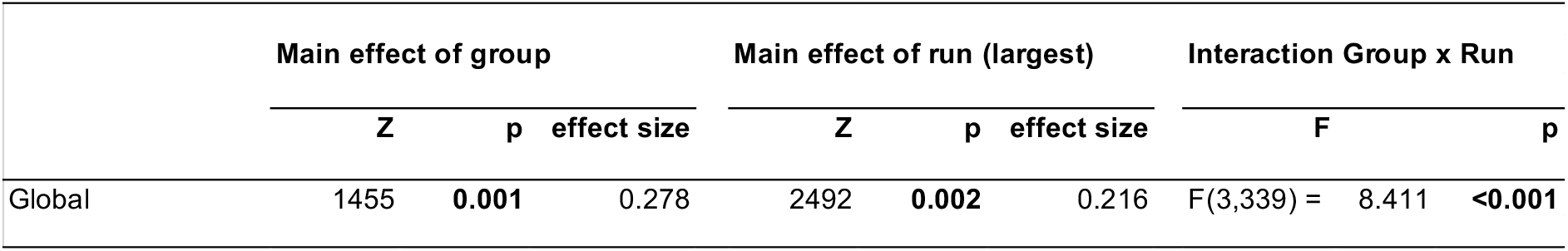
Effects of group, run, and interactions between group and run, on global VAR. Bold font indicates statistical significance

|  | Main effect of group |  |  | Main effect of run (largest) |  |  | Interaction Group x Run |  |
| --- | --- | --- | --- | --- | --- | --- | --- | --- |
|  | Z | p | effect size | Z | p | effect size | F | p |
| Global | 1455 | **0.001** | 0.278 | 2492 | **0.002** | 0.216 | F(3,339) = 8.411 | **<0.001** |

For the CON group, the effect of run was not significant. For the NAP group however, we found significant main effects of run, *χ*^2^ = 19.16, *p*<0.001, which were driven by significant differences in VAR between RUN1 and RUN3 (*p*=0.006, *effect size*=0.215), and between RUN1 and RUN4 (*p*=0.002, *effect size*=0.216).

Additionally, we found significant main effects of group which were driven by differences in VAR between CON and NAP in RUN1 (*p*=0.001, *effect size*=0.278) and RUN2 (*p*=0.002, *effect size*=0.263). As the effect size between groups in RUN1 and RUN2 were greater than the largest effect size between any pair of runs (Table 3), we inferred that metastability as measured with VAR differs between CON and NAP in RUN1 and RUN2. For the Cobre dataset, a permutation t-test for global VAR found a statistically significant difference between CON and NAP *t*(126)=-4.17, *p*<0.001 for global VAR.

**Table 3.** Effects of group, run, and interactions between group and run, on local VAR in modes *ψ*_1_,*ψ*_2_,*ψ*_3_,*ψ*_4_,*ψ*_5_. Bold font indicates statistical significance.

| Mode of Interest | Main effect of group |  |  | Main effect of run (largest) |  |  | Interaction Group x Run |  |
| --- | --- | --- | --- | --- | --- | --- | --- | --- |
|  | Z | p | effect size | Z | p | effect size | F | p |
| Mode 1 | 1434 | **0.001** | 0.287 | 2377 | **0.002** | 0.201 | F(3,339) = 8.629 | **<0.001** |
| Mode 2 | 1453 | **0.001** | 0.279 | 2322 | **0.025** | 0.168 | F(3,339) = 7.970 | **<0.001** |
| Mode 3 | 1548 | **0.005** | 0.242 | 2568 | **<0.001** | 0.278 | F(3,339) = 6.395 | **<0.001** |
| Mode 4 | 1145 | **<0.001** | 0.399 | 2851 | **<0.001** | 0.351 | F(3,339) = 20.250 | **<0.001** |
| Mode 5 | 29266 | **0.002** | 0.134 |  |  |  | F(3,339) = 2.005 | 0.113 |

### Local metastability in the spatiotemporal modes

While global VAR reflects the average VAR across the modes, it is also of interest to assess the local VAR within the modes.

For the HCPEP dataset we first investigated group (CON, NAP), run (RUN1, RUN2, RUN3, RUN4), and interactions between group and run, on global VAR for each mode *ψ*_1_, *ψ*_2_, *ψ*_3_, *ψ*_4_, *ψ*_5_. Using a 2×4 non-parametric ANOVA with the Aligned Rank Transform (ART) [55,56], we found significant interactions between group and run (Table 3).

In the CON group, we found significant main effects of run in *ψ*_4_, *χ*^2^ = 11.33, *p*=0.010. In the NAP group, we found significant main effects of run in *ψ*_1_, *χ*^2^ = 18.88, *p*<0.001, in *ψ*_2_, for *χ*^2^ = 10.60, *p*=0.014, in *ψ*_3_, *χ*^2^ = 20.12, *p*<0.001, and in *ψ*_4_, *χ*^2^ = 49.800, *p*<0.001. The drivers for these effects and the associated effect sizes are detailed in S3 Supplementary Information. The largest main effects of run are shown in Table 3.

Moreover, we found significant main effects of group in modes *ψ*_1_, *ψ*_2_, *ψ*_3_, and *ψ*_4_. The effect sizes of these differences were compared to the largest effect size between any pair of runs for that mode (Table 3). We thus found significant group differences for *ψ*_1_ in RUN1 (*p*=0.007, *effect size*=0.234) and RUN2 (*p*=0.001, *effect size*=0.287), *ψ*_2_ in RUN1 (*p*=0.003, *effect size*=0.258) and RUN2 (*p*=0.001, *effect size*=0.279), and in *ψ*_4_ in RUN1 (*p*<0, *effect size*=0.396, *moderate*) and RUN2 (*p*=0.001, *effect size*=0.399, *moderate*). We found a significant main effect of group for *ψ*_5_, *p*=0.002, *effect size*=0.134.

We thus inferred that mode VAR differed between CON and NAP in *ψ*_1_, *ψ*_2_, *ψ*_4_, and *ψ*_5_in RUN1 and RUN2, and in *ψ*_5_in all runs. For Cobre, we found statistically significant differences in mode VAR in all modes. (specifically, *ψ*_1_ *t*(125)=-3.423, *p*=0.003, *ψ*_2_ *t*(128)=-3.309, *p*=0.007, *ψ*_3_ *t*(124)=-3.584, *p*=0.002, *ψ*_4_ *t*(125)=-4.302, *p*<0.001, and *ψ*_5_*t*(128)=-4.745, *p*<0.001). Complete statistical details for global and local VAR statistics can be found in S3 Supplementary Information. Fig 4 shows the datasets with the most significant differences in mode VAR between groups.

**Fig 4.**
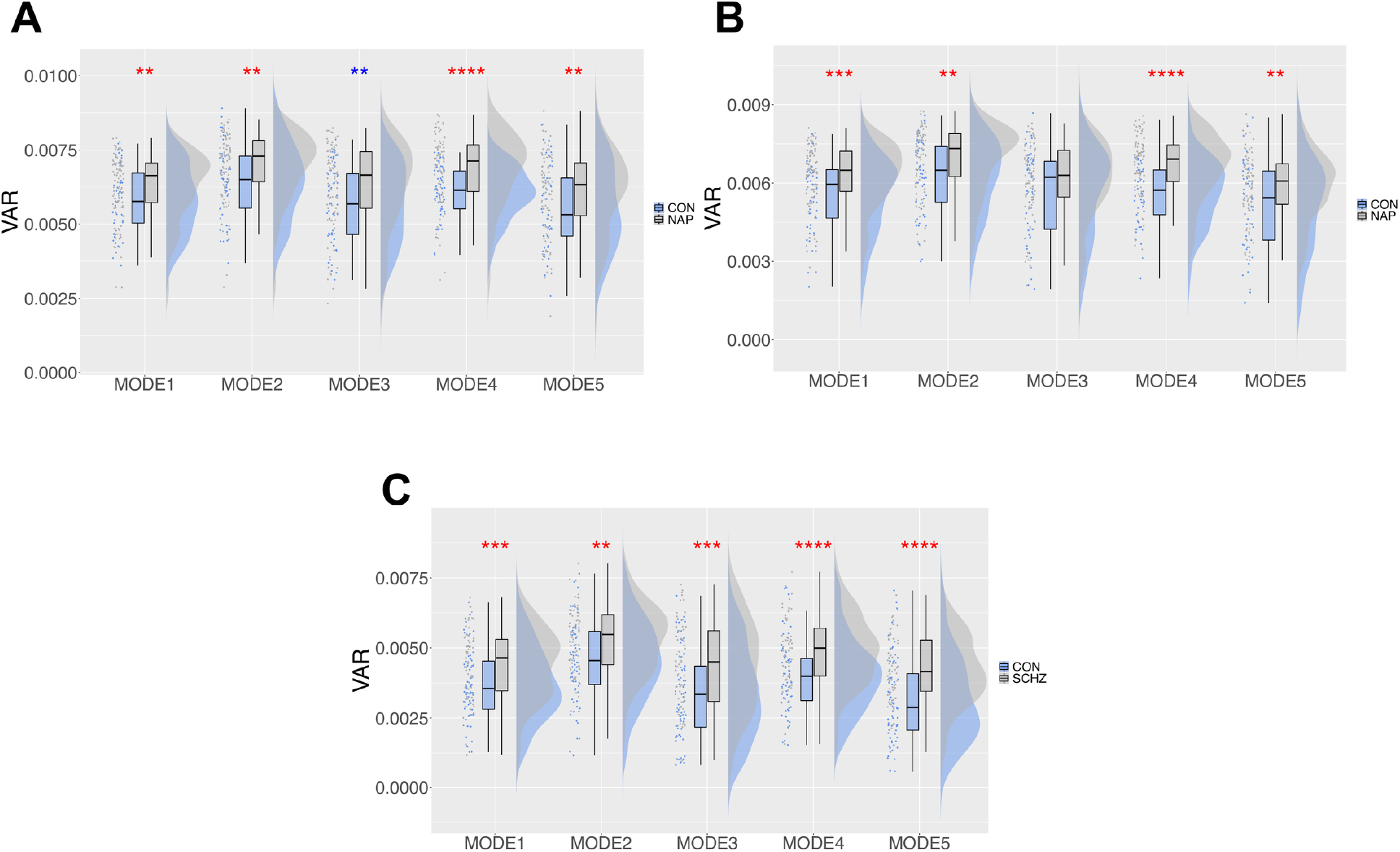
Most significant group differences in local VAR in the modes for HCPEP and Cobre datasets. Raincloud plots show from left to right the raw data, boxplots showing the median, upper and lower quartiles, upper and lower extremes, and the distributions of the raw data. **A)** HCPEP RUN1. **B)** HCPEP RUN2. **C)** Cobre dataset. *=0.05, **=0.01, ***=0.001, ****<0.001. Red * effect size between groups greater than effect size between runs. Blue * effect size between groups less than largest effect size between runs.

### Relationship with neuropsychological processes

Based on the group-level results, and the results from our basal ganglia analysis, we now highlight the differences between CON and NAP in HCPEP for Mode *ψ*_4_ in RUN2, and CON and SCHZ in Cobre for Mode *ψ*_4_. To do so we compared the connectograms for each node and the associated behavioral topics from Neurosynth meta-analysis [58]. For the meta-analysis, we applied reverse inference to gain insights into potential behavior-relevant differences between cases and controls based on their spatiotemporal modes. Following the approach of [59], we used *t*=130 terms, ranging from umbrella terms (attention and emotion) to specific cognitive processes (visual attention and episodic memory), behaviors (eating and sleep) and emotional states (fear and anxiety). The coordinates reported by Neurosynth were parcellated into 116 cortical, subcortical, and cerebellar regions. The probabilistic measure reported by Neurosynth can be interpreted as a quantitative representation of how regional fluctuations in activity are related to psychological processes. We present the comparison in Fig 5.

**Fig 5.**
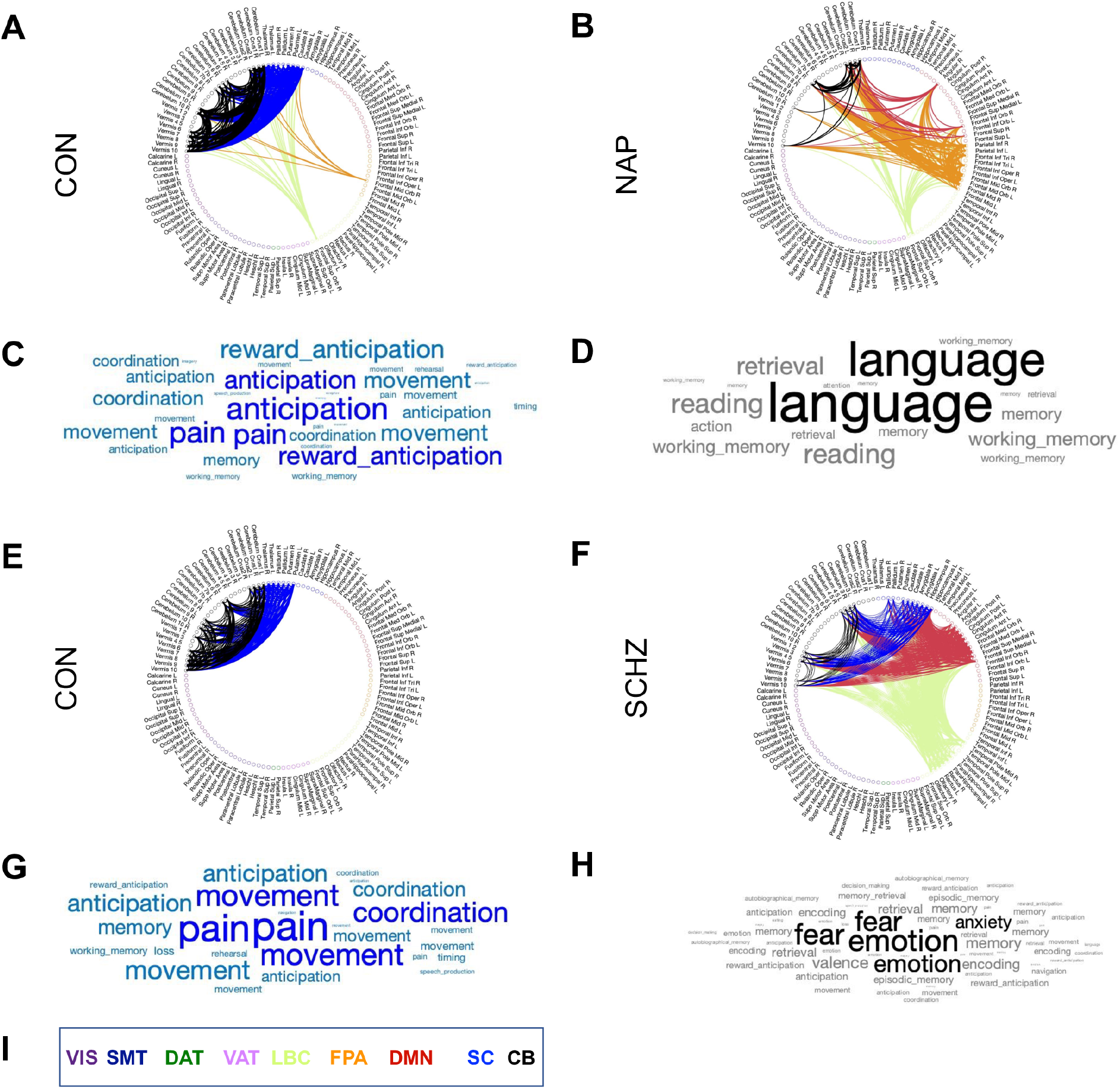
Connectograms and word clouds for Mode *ψ*_4_ in RUN2. **A)** Group-level FC in Mode *ψ*_*k*_ for HCPEP controls. **B)** Group-level FC in Mode *ψ*_*k*_ for HCPEP Non-affective psychosis. **C)** The word cloud presents the top terms derived from Neurosynth using reverse inference for the regions in Mode *ψ*_*k*_ for HCPEP controls. Word size represents the strength of the probabilistic association of the term to the regions. **D)** Top terms for Mode *ψ*_*k*_ in HCPEP Non-affective psychosis. **E)** Group-wide FC in Mode *ψ*_*k*_ for Cobre controls. **F)** Group-wide FC in Mode *ψ*_*k*_ for Cobre Schizophrenia. **G)** Top terms for Mode *ψ*_*k*_ in Cobre controls. **H)** Top terms for Mode *ψ*_*k*_ in Cobre Schizophrenia. **I)** Color coded legend for the Yeo resting-state networks, subcortical and cerebellar regions. VIS, Visual; SMT, Somatomotor; DAT, Dorsal attention; VAT, Ventral attention; LBC, Limbic; FPA, Frontal parietal; DMN, Default mode network; SC, Subcortical; CB, Cerebellar.

We see from the meta-analytical terms in HCPEP that there is an absence of anticipation and reward anticipation in the NAP group compared with the CON group. In Cobre, fear, emotion, and anxiety are present in the SCHZ group but absent in the CON group.

### Global and local metastability – individual-level neuromechanistic biomarkers of schizophrenia

Our group-level results indicated that differences in VAR across groups were statistically significant in some modes, with effect sizes being small to moderate in HCPEP, and moderate to large in Cobre (S5 Table). We therefore decided to investigate the capability of these differences to classify subjects into their relevant groups. As VAR in Mode *ψ*_4_ showed very large significant differences between groups in both HCPAP and Cobre, we decided to use this metric as an a-priori feature in a machine learning classifier.

Briefly, we used a naïve Bayes classifier with repeated k-fold cross validation (*k*=10, *repetitions*=20) on balanced samples for training and cross-validation in each dataset (HCPEP: 4 runs, Cobre: 1 run). We then tested each classifier on an out-of-sample dataset, that is trained on HCPEP, tested on Cobre, or trained on Cobre, tested on HCPEP (Table 4). HCPEP RUN2 performed best as measured by AUC when used as the training sample for out-of-sample testing in Cobre, and as the out-of-sample test for the classifier trained in Cobre. This implies that VAR in RUN2 captures best the feature that discriminates CON from NAP and SCHZ.

**Table 4.** Results of out of sample testing for each HCPEP run.

| Train | Test | AUC | B. Accuracy | Sensitivity | Specificity | p-value |
| --- | --- | --- | --- | --- | --- | --- |
| RUN1 | Cobre | 0.37 | 0.38 | 0.17 | 0.60 | 0.004 |
| RUN2 | Cobre | 0.71 | 0.58 | 0.19 | 0.96 | 0.030 |
| RUN3 | Cobre | 0.59 | 0.55 | 0.66 | 0.43 | 0.058 |
| RUN4 | Cobre | 0.57 | 0.56 | 0.87 | 0.25 | 0.049 |
| Cobre | RUN1 | 0.74 | 0.50 | 0.96 | 0.04 | 0.080 |
| Cobre | RUN2 | 0.76 | 0.57 | 0.93 | 0.21 | 0.039 |
| Cobre | RUN3 | 0.52 | 0.51 | 0.96 | 0.06 | 0.080 |
| Cobre | RUN4 | 0.40 | 0.51 | 0.93 | 0.09 | 0.080 |

HCPEP RUN2 was chosen as the training sample for external validation in Cobre, and as the validation sample for classifier trained in Cobre.

We found that although the HCPEP classifier performed better than the Cobre classifier in cross-validation (HCPEP:*AUC*=0.73, p<0.001, Cobre:*AUC*=0.71, p=0.007), the Cobre classifier performed better for out-of-sample testing (HCPEP:*AUC*=0.71, p=0.03, Cobre:*AUC*=0.76, p=0.039) as illustrated in Fig 6.

**Fig 6.**
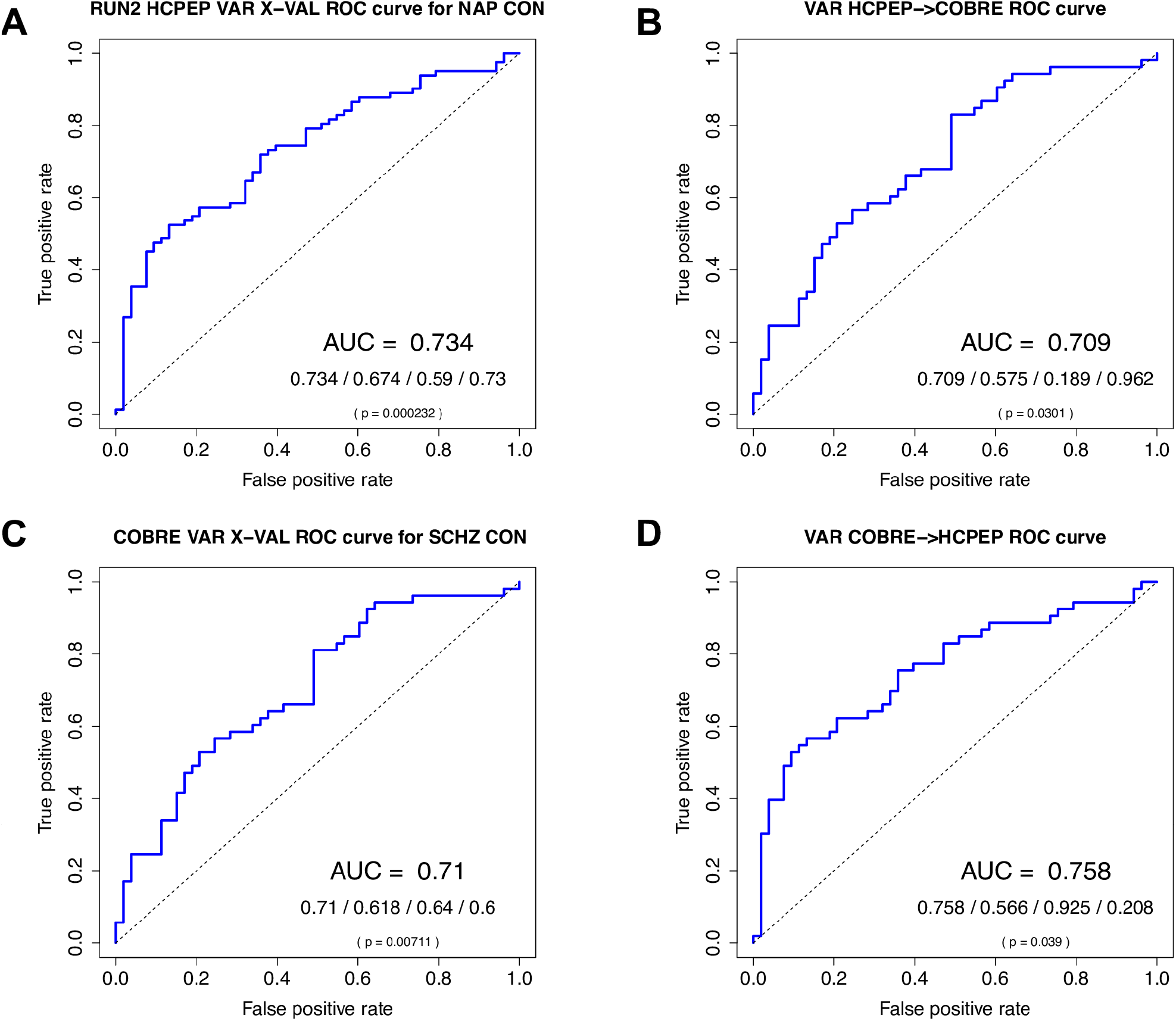
naïve Bayes classifier results for discriminating cases from controls using a single a-priori feature VAR in Mode *ψ*_4_. **A)** Results for HCPEP model trained and cross-validated in RUN 2. **B)** Results for HCPEP model trained and cross-validated in RUN 2 and tested in Cobre. **C)** Results for Cobre model trained and cross-validated. **D)** Results for Cobre model trained and cross-validated in Cobre and tested in HCPEP RUN 2. Auc/Balanced accuracy/Sensitivity/Specificity; p value calculated from the binomial distribution. AUC, area under receiver operating characteristic curve.

## Discussion

In this study we set out to assess the face validity of metastability as a neuromechanistic biomarker of schizophrenia. Our results provide preliminary evidence to support the premise that metastability measures dysfunctional connectivity in schizophrenia from 3 complementary perspectives.

First, we found statistically significant differences in group-level metastability between healthy controls and subjects with schizophrenia. Effect sizes were negligible to small (*d*=-0.16 to *d*=0.36) for early disorder subjects (NAP group) and moderate to large (*d*=-0.58 to *d*=-0.82) for subjects with established schizophrenia (SCHZ group). Previous discrimination analysis on the same Cobre dataset using a distance measure between patterns of instantaneous phase synchrony reported a moderate effect size (*d*=0.67) [28], as did another study using the mean probability of dwell time in a global state (Hedge’s *g*=0.73) [60]. In contrast, one study reported significantly lower effect sizes (*d*=0.06 to *d*=0.31) using measures of metastability in its original form, and using measures of between-network FC (*d*=0.04 to *d*=0.52) [61]. Although there are many studies that assess group-level differences in dFC, few report effect size. Therefore, limited to this small comparison, we consider that metastability, when calculated as the mean variance of instantaneous phase-locking, performs better than alternative group-level metrics reported in the literature.

Second, group-level differences in metastability (as measured with VAR) revealed group-level differences in dFC for both early and established schizophrenia. Specifically, intermittent functional disconnectivity was found for bilateral caudate, putamen left, and bilateral thalamus in early schizophrenia. The caudate and putamen are part of the dorsal striatum which is a key component in the basal ganglia. Fig 7 shows a very simple scheme of basal ganglia connectivity with the thalamus and cortex highlighting the substantia nigra pars compacta (SNc) which is the source of the neurotransmitter dopamine.

**Fig 7.**
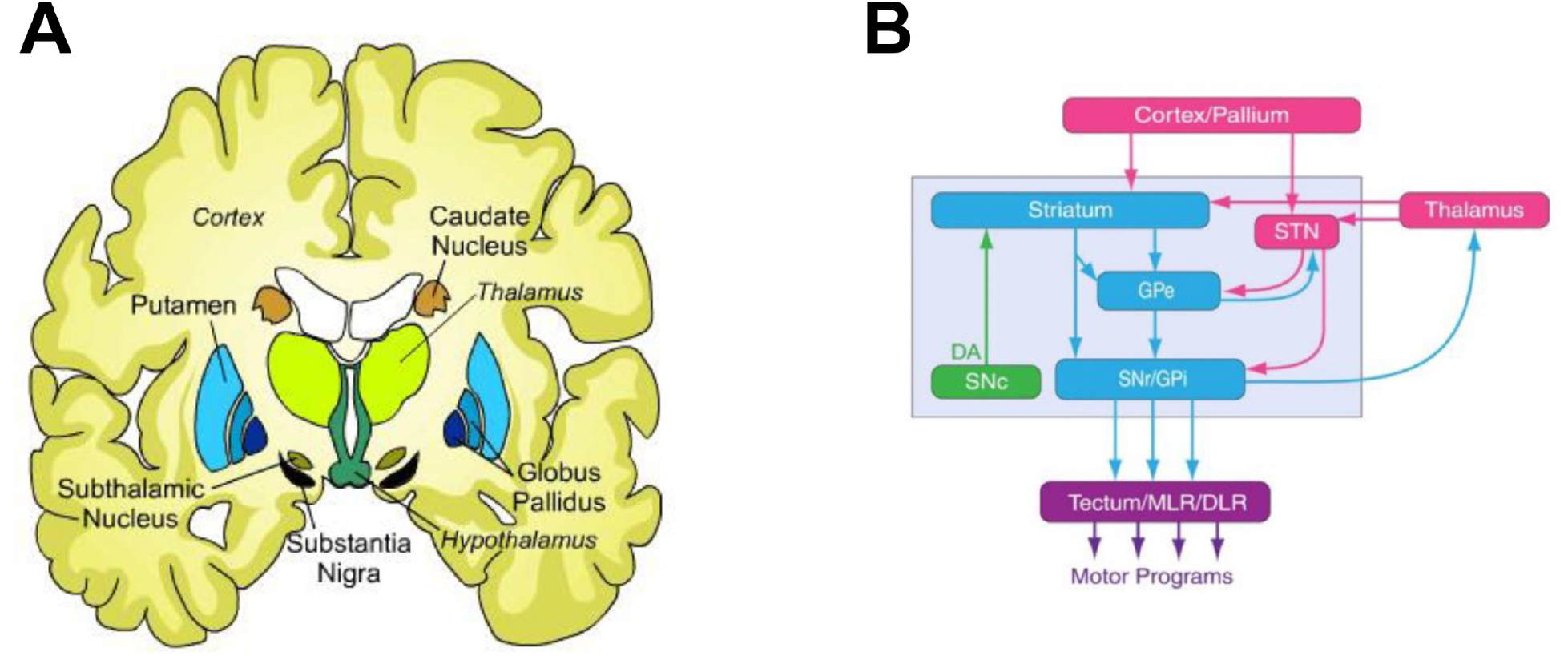
Simple scheme of basal ganglia connectivity. **A)** Location of the basal ganglia in an axial cartoon view of the brain. **B)** Basal ganglia connectivity. Arrows indicate direction of connectivity. Glutamatergic (Glu) structures are shown in rose, GABAergic nuclei are shown in cyan, and the dopaminergic (DA) nucleus is shown in green. STN, subthalamic nucleus; SNC, substantia nigra pars compacta; GPe, global pallidus external; GPi, global pallidus internal; SNr, Substantia nigra; MLR, midbrain locomotor region; diencephalon locomotor region.

Elevated dopamine synthesis and storage have been implicated in the pathophysiology of schizophrenia [62]. Hyperactivity of the substantia nigra was found to be associated with decreased prefrontal FC with basal ganglia regions in schizophrenia subjects during a working memory task [8]. In resting-state fMRI increased functional integration in the caudate and decreased FC with the prefrontal and cerebellar regions was found in subjects with schizophrenia [7]. Interestingly, striatal connectivity indices have been used to identify treatment response in first episode psychosis subjects, with higher indices associated with non-responders and lower indices associated with responders [63], which supports the hypothesis that non-responders do not possess elevated striatal dopamine synthesis capacity [64]. These findings from the literature provide evidence that that our neuromechanistic biomarker is relevant in the pathophysiology of schizophrenia.

Third, using metastability as a single a-priori feature achieved classification performance in the range of previously published studies (see Table 5). Using the Cobre dataset, one study reported quite high levels of accuracy [65] in comparison to our study, and that of Morgan et al. [66]. However, it appears that the authors did not remove cases with significant frame-wise displacement which could explain the discrepancy.

**Table 5.**
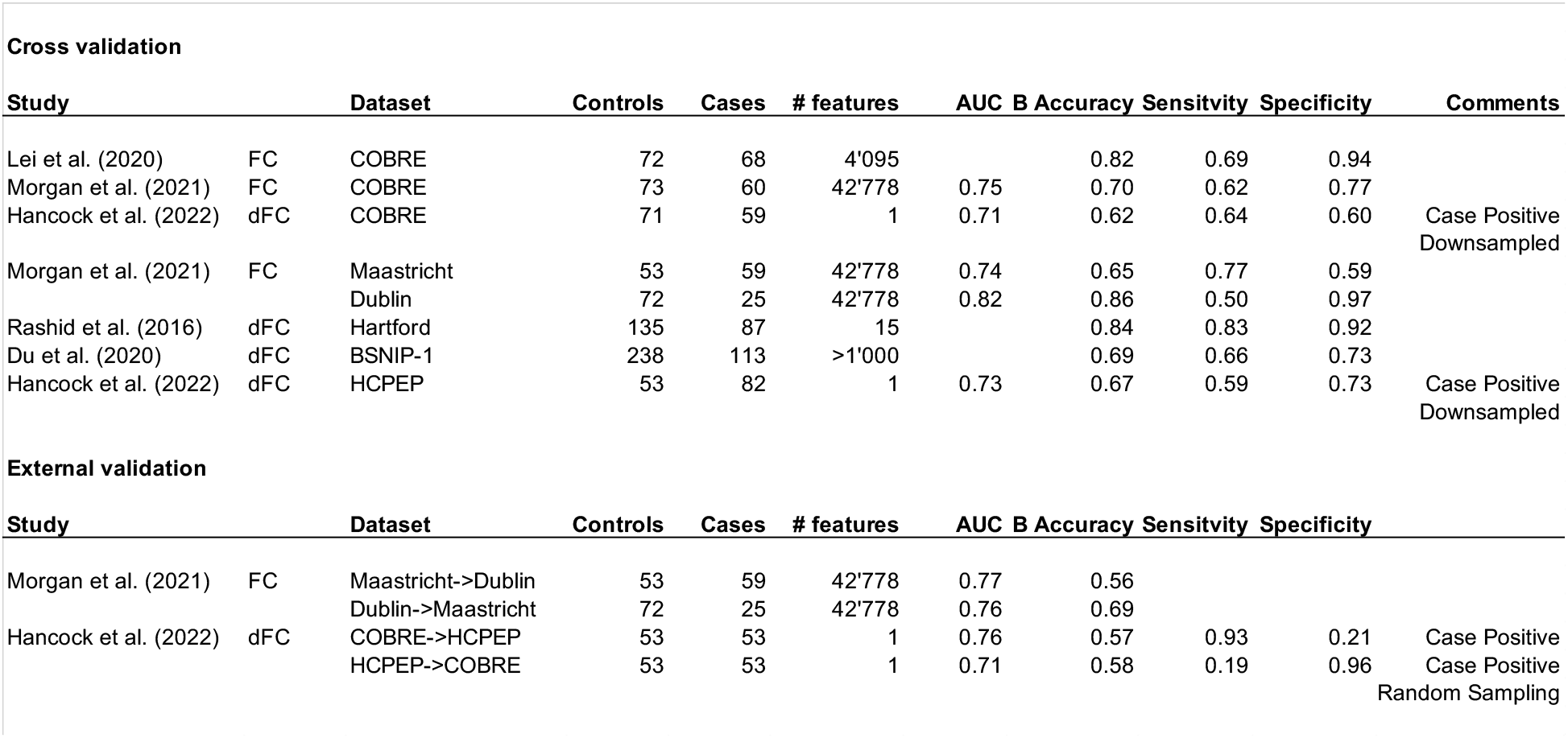
Comparison of classifier performance using FC and dFC. Case positive indicates that either NAP or SCHZ was taken as the positive class for the classifier. Down-sampled indicates that the lack of balance between classes was rectified with random down-sampling. Random sampling indicates that a specific number of samples were randomly chosen to allow balanced classes for external cross-validation. Blank cells indicate that the information was not available in the study manuscript.

When considering classification performance in different datasets, it appears that our classifier did not perform as well as the one from Morgan et al. [66] in the Dublin dataset, or with the one from Rashid et al. [18] in the Hartford dataset. However, in both cases the classes were not balanced (Dublin, cases:controls = 25:75, Hartford, cases:controls = 87:135) and there was no evidence that this was taken into consideration when reporting the performance, which may explain the discrepancy in the results.

We note that our cross-validation performance is comparable to the cross-validation performance in the other studies. Given these comparisons, we consider that our classifier had similar performance to those reported in the literature for cross-validation.

When we compare external validation of our classifier to that of Morgan et al. [66], we see that performance is similar with the same caveat pertaining to the Dublin dataset. We therefore consider that our cross-dataset analysis based on a single a-priori feature of metastability (as measured with VAR) performs similarly to one in the literature based on over 40’000 features in FC.

It is interesting to note that the HCPEP->Cobre external validation returned very high specificity whilst the Cobre->HCPEP external validation returned very high sensitivity. This may reflect that the cases in HCPEP are in the early stages of schizophrenia whilst the cases in Cobre are in a well-established stage of schizophrenia. It appears that disruptions in connectivity in early psychosis are not sufficient to distinguish SCHZ from CON. However, disruptions in SCHZ are sufficient to distinguish NAP from CON. This seems to imply that the disruptions in early psychosis are a subset of those in established schizophrenia.

Our three complementary perspectives of group-level discrimination, individual-level classification, and pathophysiological relevance, provide preliminary evidence for the face validation of metastability as a neuromechanistic biomarker of schizophrenia.

There are several limitations that should be considered when evaluating the findings from this study. First, we used a novel proxy for metastability. Although the concept of metastability is generally accepted, its operationalization takes a number of forms from the entropy of spectral density [31], to the variability in spatial coherence [67], and to the most commonly used form, the standard deviation of the Kuramoto order parameter (phase coherence) [57]. We chose the mean variance of instantaneous phase-locking as an alternative proxy for metastability based on the theory of Synergetics [45] and recent generalization of the Haken-Kelso-Bunz (HKB) model to multiple oscillators [46], which exhibits stable antiphase synchronization [47]. See Fig 8 reproduced with permission from [47]. It should be noted that the generalized HKB model reduces to the Kuramoto model when second-order coupling is removed (i.e. B_ij_ = 0), and so can be seen as an extension of the Kuramoto model as in [68–70].

**Fig 8.**
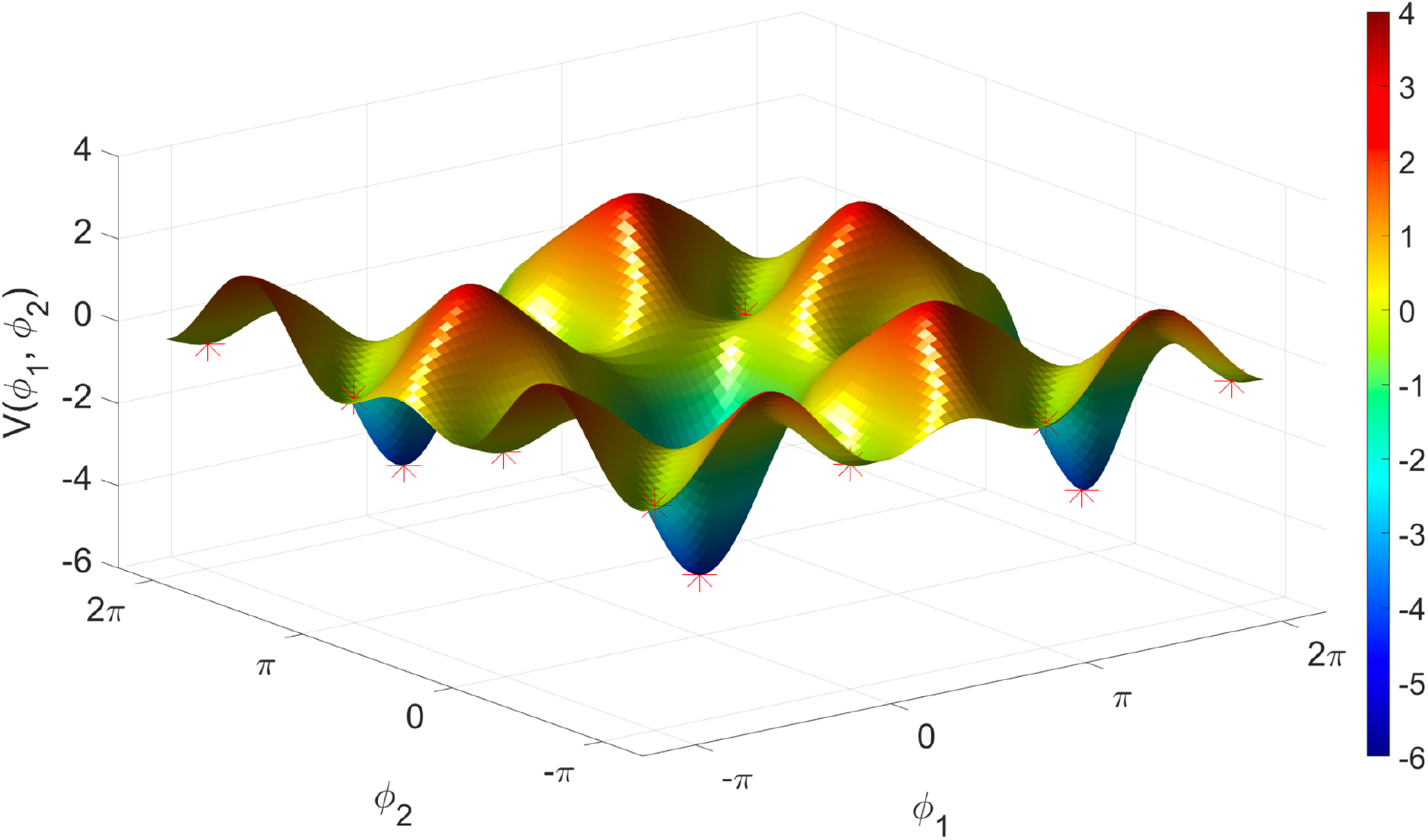
Attractor landscape for the extended HKB model of multi-adic coordination. A plot of the relative phase potential function landscape for Aij = 2Bij = 1 for each i, j. Note the many valleys (marked with red asterisks) in which an oscillator moving around in this landscape will become trapped. These valleys are the local minima corresponding to the coordination states. There are two types of valleys in this landscape: in-phase valleys, which have relatively very deep and wide basins of attraction, and antiphase valleys, which are narrower and shallower, reflecting the fact that the in-phase state is more stable than the antiphase state. Each of these valleys is separated by a distance of Π, and repeats infinitely on the potential surface in a 2 Π -periodic pattern. A, B, effective coupling parameters; i,j, i^th^ and j^th^ oscillator. Reproduced with permission from [47].

The generalized HKB model may explain the phase-locking behavior we illustrated in Fig 1 including mono-stability, bi-stability, switching (‘sans switch’) [71], and chimeras [57]. We have used a phenomenological understanding of the generalized HKB model to propose the mean variation of instantaneous phase-locking as a new proxy measure for metastability. In future work we need to perform a more thorough theoretical investigation of the phenomenon of metastability, complemented with a computational model that predicts empirical findings.

Second, from the perspective of alternative dFC approaches and pipelines, we did not perform global signal, white matter or cerebral spinal fluid regression. From a complexity science perspective [34], one cannot explore any subsystem of a complex system such as the brain in isolation, and accumulating evidence points to contributions other than neuronal to the fMRI signal [72–74]. As in [42], we defined communities of oscillators directly from the phase-locking data and not from intrinsic connectivity networks [36] nor predefined templates [61]. This allows regions to participate in multiple communities reflecting transient coalitions between the regions as evidenced by spatial overlap between networks [75,76].

Third, the switching behavior observed in the phase-locking behavior in Fig 1 may appear to be artifactual. In LEiDA the leading eigenvector time-series is smoothed through a technique called “half-switching”. We reproduced the time-series for one subject without this smoothing and compared the results to the smoothed version. As may be seen in S5 Fig switching also occurs in the non-smoothed version, but with higher frequency than in the smoothed version.

However, since this smoothing was applied to all subjects, it does not affect the results, but may impact the ability to compare results with those obtained with alternative dFC approaches.

## Conclusion

This study claims face validity of metastability as a neuromechanistic biomarker of schizophrenia based on group-level discrimination, individual-level classification, and pathophysiological relevance, congruent with published literature. While diagnostic biomarkers of schizophrenia — such as metastability — may still have limited clinical utility, they can provide mechanistic insights for the discovery of prognostic biomarkers that could support treatment decisions. For example, the ability to identify treatment resistance or transition likelihood from high risk to first episode psychosis would address a real clinical need. Developing a deeper understanding of metastability may one day help us to gain sufficient mechanistic insight into the disconnection phenomenon of schizophrenia, which may lead in turn into the development of such effective biomarkers.

## Materials and methods

### Participants

#### HCPEP

Healthy controls (CON, *n* = 53) and non-affective psychosis (NAP, *n* = 82) participants were scanned at one of four sites (Indiana University, Beth Israel Deaconess Medical Center – Massachusetts Mental Health Center, McLean Hospital and Massachusetts General Hospital) as part of the Human Connectome Project-Early Psychosis (Principal Investigators: Shenton, Martha; Breier, Alan; U01MH109977-01, HCP-EP; <u>doi:10.15154/1524263</u> https://nda.nih.gov/edit_collection.html?id=2914) with funding from the National Institute of Mental Health (NIMH). A Data Use Certification (DUC) is required to access the HCPEP on the NIMH Data Archive (NDA).

NAP participants met DSM-5 criteria for schizophrenia, schizophreniform, schizoaffective, psychosis NOS, delusional disorder, or brief psychotic disorder with onset within the past five years prior to study entry. Additional inclusion/exclusion criteria may be found in http://www.humanconnectome.org/storage/app/media/documentation/data_release/HCP-EP_Release_1.0_Manual.pdf. See Table 6 for group demographics.

**Table 6.** Demographic characteristics of participant groups

| Characteristics | HCP EP |  | COBRE |  |
| --- | --- | --- | --- | --- |
|  | HCS (n=53) | NAPs (n=82) | HCS (n=72) | SCHZs (n=72) |
| Age (years) | 24.85 ± 4.15 | 23.42 ± 3.57 | 35.88 ± 11.74 | 37.89 ± 13.86 |
| Gender (male/female) | 34/19 | 55/27 | 51/21 | 56/16 |
| Site (IU/BIDMC/MGH/MH) | 25/5/10/13 | 51/14/5/12 |  |  |
| IQ (QASI-II) (160 subjects recorded) | 116.30 ± 10.96 | 97.52 ± 17.55 |  |  |

*HCP EP = Human Connexome Project for Early Psychosis; HC = Healthy Controls; NAP = patients with non-affective psychosis; SCHZ = patients with schizophrenia; IQ = intelligence quotient*

Procedures were approved by the Partners Healthcare Human Research Committee/IRB and complied with the Declaration of Helsinki. Participants provided written informed consent, or in the case of minors, parental written consent and participant assent.

#### Cobre

Neuroimaging data was obtained from the publicly available repository Cobre (http://fcon_1000.projects.nitrc.org/indi/retro/cobre.html) preprocessed with NIAK 0.17—lightweight release (Calhoun et al., 2012; Bellec, 2016). The neuroimaging data included preprocessed resting-state fMRI data from healthy controls (CON, *n* = 72) and schizophrenia patients (SCHZ, *n* = 72), in which participants passively stared at a fixation cross. Subject recruitment and evaluation may be found in (Aine et al., 2017). The study was approved by the institutional review board (IRB) of the University of New Mexico (UNM) and all subjects provided written informed consent. See Table 6 for group demographics.

#### Image acquisition - HCPEP

All MRI scans were acquired on Siemens MAGNETOM Prisma 3T scanners with a multiband acceleration factor of 8, and a 32/64channel head coil. Each participant underwent four scans of resting-state fMRI collected over two experimental sessions on consecutive days (two scans in each session). The four datasets are referred to as run 1 to run 4. During each scan 410 frames were acquired using a multiband sequence at 2 mm isotropic resolution with repetition time (TR) of 0.72 sec over the space of 4 min 55 secs. The two scans in each session differed only in the phase encoding direction of anterior-posterior (AP) followed by posterior-anterior (PA) on both days.

#### Image acquisition - Cobre

The resting-state fMRI data featured 150 echo planar imaging volumes obtained in 5 min, with repetition time (TR) = 2 s, echo time = 29 ms, acquisition matrix = 64×64 mm2, flip angle = 75° and voxel size = 3×3×4 mm3. The acquisition is fully described in detail in (Aine et al., 2017).

### Preprocessing

#### HCPEP

Data were pre-processed with the HCP’s minimal pre-processing pipeline, and denoising was performed by the ICA-FIX procedure (Glasser et al., 2013; Griffanti et al., 2014; Salimi-Khorshidi et al., 2014). A complete description of the pre-processing details may be found at the HCP website https://www.humanconnectome.org/software/hcp-mr-pipelines. Briefly, fMRI data was gradient-nonlinearity distortion corrected, rigidly realigned to adjust for motion, fieldmap corrected, aligned to the structural images, and then registered to MNI space with the nonlinear warping calculated from the structural images. ICA-FIX was then applied on the data to identify and remove motion and other artifacts in the time-series.

#### Cobre

The preprocessing of the fMRI data is fully described in detail in [77,78].

In brief, preprocessing included slice-timing correction, co-registration to the Montreal Neurological Institute (MNI) template and resampling of the functional volumes in the MNI space at a 6 mm isotropic resolution. We resampled the functional volumes in MNI space at a 2 mm isotropic resolution with 3dresample from AFNI [79]. Inspection of the fMRI data for each subject resulted in the exclusion of one subject whose data did not include all 150 volumes. 13 NAP subjects with framewise displacement > 0.7mm were also removed. The final dataset therefore used for the Cobre analysis included n=59 SCHZ cases and n=71 HCs.

Substantial material in the following subsections is recycled from our prior publication [42].

### Parcellation

We parcellated the pre-processed fMRI data by averaging time-courses across all voxels for each region defined in the anatomical parcellation AAL [48] considering all cortical, subcortical, and cerebellar regions, *N* = 116. We chose the AAL parcellation as subcortical and cerebellar regions are relevant in studies with psychiatric cohorts [2,7,80–82].

### Bandpass filtering

To isolate low-frequency resting-state signal fluctuations, we bandpass filtered the parcellated fMRI time-series within 0.01-0.08 Hz with a discrete Fourier transform (DST) computed using a fast Fourier transform (FFT) algorithm in MATLAB 2021b. We applied Carson’s empirical rule (Carson, 1922; Pachaud et al., 2013) on the analytical signal which was calculated using the Hilbert transform of the real signal (Gabor, 1946), to confirm non-violation of the Bedrosian theorem for our band-passed signals in both datasets (see S3 and S4 Figs).

### Functional connectivity through phase-locking

We estimated functional connectivity (FC) with the nonlinear measure of phase-locking which may be more suitable than linear measures such as Pearson correlation for analyzing complex brain dynamics. Specifically, nonlinear methods provide insight into interdependence between brain regions at both short and large time and spatial scales allowing the analysis of complex nonlinear interactions across space and time [83,84]. From a practical perspective, unlike correlation or covariance measures, phase synchronization can be estimated at the instantaneous level and does not require time-windowing. When averaged over a sufficiently long-time window, phase-locking values provide a close approximation to Pearson correlation, varying within the same range of values [50,85].

Following [50] we first calculated the analytical signal using the Hilbert transform of the real signal [86]. Then, the instantaneous phase-locking between each pair of brain regions *n* and *p* was estimated for each time-point *t* as the cosine difference of the relative phase as

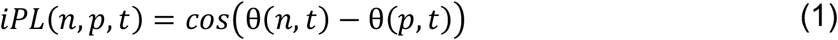

Phase-locking at a given timepoint ranges between -1 (regions in anti-phase) and 1 (regions in-phase). For each subject the resulting *iPL* was a three-dimensional tensor of size *NxNxT* where *N* is the dimension of the parcellation, and *T* is the number of timepoints in the scan.

### LEiDA – Leading Eigenvector Dynamic Analysis

To reduce the dimensionality of the phase-locking space for our dynamic analysis, we employed the Leading Eigenvector Dynamic Analysis (LEiDA) [50] method. The leading eigenvector *V*_*1*_*(t)* of each *iPL(t)* is the eigenvector with the largest magnitude eigenvalue and reflects the dominant FC (through phase-locking) pattern at time *t. V*_*1*_*(t)* is a *Nx1* vector that captures the main orientation of the fMRI signal phases over all anatomical areas. Each element in *V*_*1*_*(t)* represents the projection of the fMRI phase in each region into the leading eigenvector. When all elements of *V*_*1*_*(t)* have the same sign, this means that all fMRI phases are orientated in the same direction as *V*_*1*_*(t)* indicating a global mode governing all fMRI signals. When the elements of *V*_*1*_*(t)* have both positive and negative signs, this means that the fMRI signals have different orientations behaving like opposite anti-nodes in a standing wave. This allows us to separate the brain regions into two ‘communities’ (or poles) according to their orientation or sign, where the magnitude of each element in *V*_*1*_*(t)* indicates the strength of belonging to that community [87]. For more details and graphical representation see [51,88,89]. The outer product of *V*_*1*_*(t)* reveals the FC matrix associated with the leading eigenvector at time *t*.

### Mode extraction

To identify recurring spatiotemporal modes *ψ* or phase-locking patterns, we clustered the leading eigenvectors for each of the 12 phase-locked time-series datasets (3 conditions x 4 runs) with K-means clustering with 300 replications and up to 400 iterations for 2-10 centroids. K-means clustering returns a set of K central vectors or centroids in the form of *Nx1* vectors *V*_*c*_. As *V*_*c*_ is a mean derived variable, it may not occur in any individual subject data set. To obtain time courses related to the extracted modes *ψ*_*k*_ at each TR we assign the cluster number to which *V*_*c*_*(t)* is most similar using the cosine distance.

### Mode representation in voxel space

To obtain a visualization in voxel space of the spatial modes *V*_*c*_ we first reduced the spatial resolution of all fMRI volumes from 2mm^3^ to 10mm^3^ to obtain a reduced number of brain voxels (here *N* = 1821) to be able to compute the eigenvectors of the *NxN* phase-locking matrices. The analytic signal of each 10mm^3^ voxel was computed using the Hilbert transform, and the leading eigenvectors were obtained at each time point (with size *NxT*). Subsequently, the eigenvectors were averaged across all time instances assigned to a particular cluster, obtaining in this way, for each cluster, a *1xN* vector representative of the mean phase-locking pattern captured in voxel space.

### Mode representations as connectograms

We visualized FC as connectograms by taking the FC matrices for each mode and retaining regions that were collectively in-phase but in anti-phase with the global mode.

### Neurosynth functional associations

Probabilistic measures of the association between brain coordinates and overlapping terms from the Cognitive Atlas [90] and the Neurosynth database [58] were obtained as in [59]. The probabilistic measures were parcellated into 116 AAL regions and may be interpreted as a quantitative representation of how regional fluctuations in phase-locking are related to psychological processes. The resulting functional association matrix represents the functional relatedness of 130 terms to 116 brain regions (see S7 Table for a full list of terms).

### Metastability

Empirical metastability studies to date have used pre-defined resting-state networks (RSN) extracted with ICA [36], with network masks [61], or with functional templates [91] to represent communities of oscillators for investigation of network synchrony and metastability. In contrast, as in [42] we decided to take a purely data driven approach, using the recurrent modes extracted with K-means clustering to represent communities of oscillators. As we decided to retain 5 recurrent modes (see Results), we therefore have 5 communities of oscillators *ψ*_1_ - *ψ*_5_. Note that the AAL regions are not constrained to a single community and so the communities reflect time-varying coalitions among regions.

#### Based on phase synchrony

Synchronization was calculated as the time-average of the Kuramoto order parameter in each community, which is given by

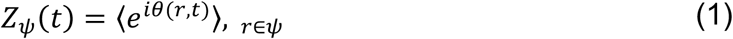

Above, *Z*_*ψ*_(*t*) is a complex value where its magnitude, and hence *SYNC*_*ψ*_= |*Z*_*ψ*_(*t*)|, provides a quantification of the degree of synchronization of the community at each time *t*, taking values between 1 (for fully synchronized systems) and 0 (for fully desynchronized systems). Metastability was calculated as the standard deviation over time of the Kuramoto order parameter in each community. The mean value of this measure across communities denoted as global metastability, represents the overall variability in the synchronization across communities.

#### Based on phase-locking

The instantaneous phase-locking between each pair of brain regions *n* and *p* was estimated for each time-point *t* as the cosine difference of the relative phase as

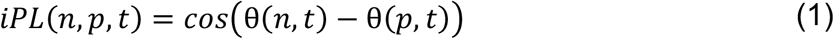

Metastability, denoted as VAR to distinguish it from metastability above, was calculated as the mean of the variance of instantaneous phase-locking over time in each community. The mean value of this measure across communities denoted as global VAR represents the overall variance in the phase-locking across communities.

### Statistical analysis

#### Interclass correlation coefficient

ICC is a relative metric that is used for test-retest reliability in measurement theory [92]. It is generally defined as the proportion of the total measured variance that can be attributed to within subject variation. As such, ICC coefficients may be low when there is little variance between subjects, that is in a homogeneous sample, or when the within-subject variance is large [93]. In this study we use the ICC forms from [94]. There are many scales for ICC, so for clarity we will use those of Landis and Koch [95]:

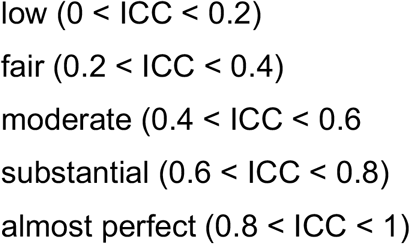

We calculated the run reliability of mode *ψ* extraction with ICC(1,1) in search of agreement rather than consistency across runs [54].

#### Parametric testing

Before performing statistical tests, we checked if the assumptions for parametric testing were met. In all cases, the assumptions were violated. The results of these tests can be found for basal ganglia in S1 Table, global metastability in S2 Table, local metastability in S3 Table, global VAR in S4 Table, local VAR in S5 Table.

#### Non-parametric ANOVA testing

We used Align rank transform (ART) [55,56] to perform multi-factor non-parametric testing with dependent groups in R (ARtool::art). We then followed the statistical testing flowchart shown in Fig 9. All results were Bonferroni corrected for multiple comparisons.

**Fig 9.**
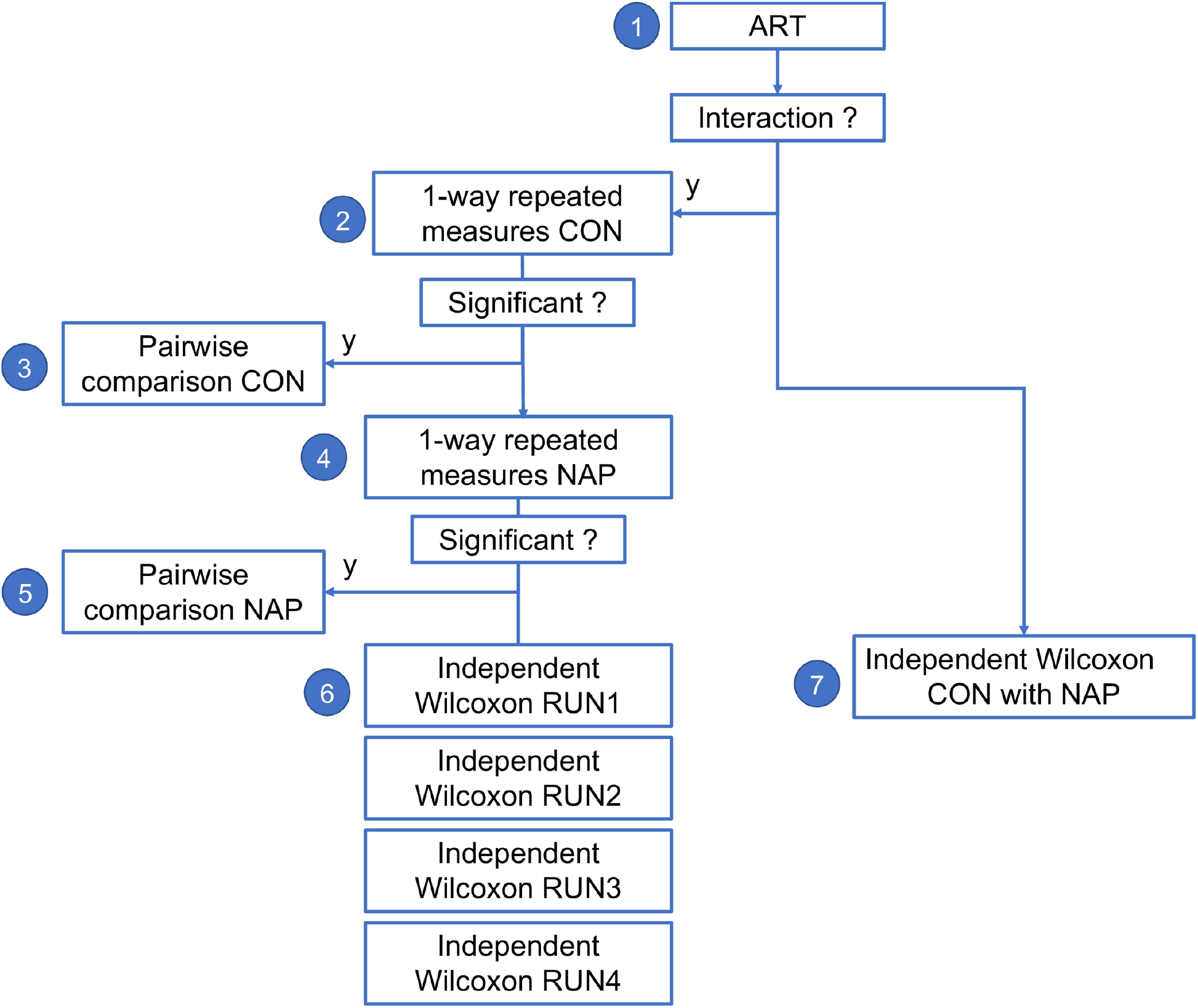
Statistical flowchart for non-parametric testing of differences between groups across runs. 1) 2×4 non-parametric ANOVA using Align rank transform (ART). 2) Friedmann repeated measures test. 3) Paired Wilcoxon test. 4) Friedmann repeated measures test. 5) Paired Wilcoxon test. 6) Independent Wilcoxon test for each run. 7) Independent Wilcoxon test across all runs.

#### Non-parametric permutation t-tests

We used permutation Welch 2 sample t-tests with *n* = 9999 Monte Carlo permutations implemented in R (MKinfer::perm.t.test) as the majority of distributions were not normally distributed when assessed with a Shapiro test.

#### Classification of condition based on metastability

Supervised machine learning algorithms were trained to classify cases and controls for each dataset independently using a single a-priori feature of metastability as measured by VAR. Classification was performed using a naïve Bayes non-linear classification model [96] in R implemented with Caret [97]. We used a naïve Bayes classifier as we had just one feature with no issue of independence. For the HCPEP datasets, we chose cross-validation over internal validation in a different run to avoid data leakage, as the same participants would have been present in both the test and validation sets [98].

In all five datasets, we assessed the generalizability of the classifier using repeated k-fold cross-validation, *k*=10, *repetitions* = 20. For the out-of-sample analysis we trained the classifier in HCPEP and tested it in Cobre; and trained the classifier in Cobre and tested it in HCPEP. For all datasets we used down-sampling to balance the classes, and for the out-of-sample analysis we randomly down-sampled both datasets to 53 to allow cross-dataset testing. We report the area under the operating characteristics curve (AUC), balanced accuracy, sensitivity, and specificity. The statistical significance of balanced accuracy was assessed with a binomial cumulative distribution function [98].

#### Software tools

Parcellation, LEiDA, ICC and metastability / VAR derivations were implemented in MATLAB [99]. Neurosynth functional associations were derived in Python 3.8.5. All other statistical analysis were performed in RStudio Team version 2022.02.3 Build 492 [100].

## Data Availability

Data were obtained from the National Institute of Mental Health (NIMH) Data Archive (NDA; study DOI: 10.15154/1524263). NDA is a collaborative informatics system created by the National Institutes of Health to provide a national resource to support and accelerate research in mental health.
The Cobre data were downloaded from the Collaborative Informatics and Neuroimaging Suite Data Exchange tool (COINS; http://coins.mrn.org/dx), and data collection was performed at the Mind Research Network and funded by a Center of Biomedical Research Excellence Grant No. 5P20RR021938/P20GM103472 from the National Institutes of Health to Dr. Vince Calhoun.

https://github.com/franhancock/Meta_Schz

## Supporting information

**S1 Fig Silhouette values for clustering solutions for 1 to 9 clusters with 2-10 modes respectively**. (**A**) HCPEP CON. (**B**) HCPEP NAP. (**C**) Cobre CON **(D)** Cobre SCHZ.

**S2 Fig Reliability of mode extraction for controls and non-affective psychosis**

**S3 Fig Non-violation of Bedrosian Theorem - HCPEP**

**S4 Fig Non-violation of Bedrosian Theorem – Cobre**

**S5 Fig Effect of smoothing on the leading eigenvector time-series**. A) Timeseries for the leading eigenvectors for one subject without smoothing. B) Timeseries for the leading eigenvector for the same subject with half-switch smoothing. The blue asterixis indicate that half-switching occurred.

**S1 Table Assumption test results for contribution of basal ganglia regions FC in the HCPEP dataset**. We assessed the normality of the distribution of contribution with a Shapiro-Wilk test, equivalence of variance with a Levine test, and effect size with Cohen’s D test.

**S2 Table Assumption test results for global META in the HCPEP and Cobre datasets**. We assessed the normality of the distribution of META with a Shapiro-Wilk test, equivalence of variance with a Levine test, and effect size with Cohen’s D test.

**S3 Table. Assumption test results for metastability in the modes in the HCPEP and Cobre datasets**. We assessed the normality of the distribution of mode META with a Shapiro-Wilk test, equivalence of variance with a Levine test, and effect size with Cohen’s D test.

**S4 Table. Assumption test results for global VAR in the HCPEP and Cobre datasets**. We assessed the normality of the distribution of META with a Shapiro-Wilk test, equivalence of variance with a Levine test, and effect size with Cohen’s D test.

**S5 Table 7. Assumption test results for mode VAR in the HCPEP and Cobre datasets**. We assessed the normality of the distribution of VAR in each mode with a Shapiro-Wilk test, equivalence of variance with a Levine test, and effect size with Cohen’s D test.

**S6 Table. Neurosynth terms**

**S1 Supplementary Information**. Results from statistical tests for differences in basal ganglia connectivity between HC and NAP in the HCPEP dataset

**S2 Supplementary Information** Results from statistical tests for differences in mode META between HC and NAP in the HCPEP dataset, and HC and SCHZ in the Cobre dataset

**S3 Supplementary Information** Results from statistical tests for differences in mode VAR between HC and NAP in the HCPEP dataset, and HC and SCHZ in the Cobre dataset

**S1 Supplementary Text** Analysis of coherence based Metastability

### Acknowledgements

Data were obtained from the National Institute of Mental Health (NIMH) Data Archive (NDA; study DOI: 10.15154/1524263). NDA is a collaborative informatics system created by the National Institutes of Health to provide a national resource to support and accelerate research in mental health. The Cobre data were downloaded from the Collaborative Informatics and Neuroimaging Suite Data Exchange tool (COINS; http://coins.mrn.org/dx), and data collection was performed at the Mind Research Network and funded by a Center of Biomedical Research Excellence Grant No. 5P20RR021938/P20GM103472 from the National Institutes of Health to Dr. Vince Calhoun. All authors report no financial interests or potential conflicts of interest.

## Author Contributions

**Conceptualization:** Fran Hancock

**Data curation:** Robert A. McCutcheon

**Formal analysis:** Fran Hancock

**Investigation:** Fran Hancock

**Methodology:** Fran Hancock, Joana Cabral

**Software:** Fran Hancock

**Supervision:** Federico E. Turkheimer, Ottavia Dipasquale

**Validation:** Fran Hancock

**Visualization:** Fran Hancock

**Writing – original draft:** Fran Hancock, Fernando E. Rosas, Robert A. McCutcheon

**Writing – review and editing:** Fran Hancock, Fernando E. Rosas, Robert A. McCutcheon, Joana Cabral, Ottavia Dipasquale, Federico E. Turkheimer

