## Supplementary Material for "Metastability as a neuromechanistic biomarker of schizophrenia pathology"

### Supplementary Text S1

### Coherence based metastability

#### Global and local metastability – group-level neuromechanistic biomarkers of schizophrenia

We calculated global and local metastability to compare group-level dynamics between case-control groups as the standard deviation of instantaneous phase synchrony as in [1]–[4]. Metastability measures the balance between functional integration and functional segregation, has been found to be stable across multiple acquisitions in healthy young adults [1], and is perhaps uniquely suitable for assessing schizophrenia. We calculated global and local metastability within each mode and for each run.

##### Global metastability

For the HCPEP dataset we first investigated group (CON, NAP), run (RUN1, RUN2, RUN3, RUN4), and group x run effects on global META. Using a 2x4 non-parametric ANOVA with the Aligned Rank Transform (ART) [53], [54], we found a significant interaction $F(3,399)=3.192$, $p=0.024$ for group x run.

##### Main effects of run

The main effect of run was not significant for either the CON or the NAP group.

#### Main effects of group

We found significant main effects of group which were driven by differences in META between CON and NAP in RUN4 (*p*=0.019, *effect size*= 0.202). See Table 1 for complete results.

**Table 1. Non-parametric statistical tests for differences in global META between CON and NAP across runs.**

| | META | | | | | | | | | | |
|---|---|---|---|---|---|---|---|---|---|---|---|
| **Step 1** | **2x4 non-parametric ANOVA using ART** | | | | | | | | | | |
| | | **Term** | **F** | **Df** | **Df.res** | **Pr(>F)** | | | | | |
| | COND | COND | 0.235 | 1 | 133 | 0.629 | | | | | |
| | RUN | RUN | 1.302 | 3 | 399 | 0.273 | | | | | |
| | COND:RUN | COND:RUN | 3.192 | 3 | 399 | 0.024 | | | | | |
| **Step 2** | **CON:1x4 repeated measures non-parametric ANOVA using Friedmann test** | | | | | | | | | | |
| | **chi-squared** | **df** | **p.value** | **method** | **data** | | | | | | |
| | 0.872 | 3 | 0.832 | Friedman | META and RUN and SUB | | | | | | |
| **Step 4** | **NAP:1x4 repeated measures non-parametric ANOVA using Friedmann test** | | | | | | | | | | |
| | **chi-squared** | **df** | **p.value** | **method** | **data** | | | | | | |
| | 6.673 | 3 | 0.083 | Friedman | META and RUN and SUB | | | | | | |
| **Step 6** | **Non-parametric Wilcox test between CON and NAP for each run** | | | | | | | | | | |
| | | **.y.** | **group1** | **group2** | **n1** | **n2** | **statistic** | **p** | **p.signif** | **effsize** | **magnitude** |
| | RUN1 | META | CON | NAP | 53 | 82 | 2082 | 0.683 | ns | 0.035 | small |
| | RUN2 | META | CON | NAP | 53 | 82 | 2131 | 0.852 | ns | 0.016 | small |
| | RUN3 | META | CON | NAP | 53 | 82 | 2003 | 0.445 | ns | 0.066 | small |
| | RUN4 | META | CON | NAP | 53 | 82 | 2694 | 0.019 | * | 0.202 | small |

ART, Aligned Rank Transform; CON, controls; NAP, non-affective psychosis

For the Cobre dataset, we found a statistically significant difference $t(126)=-3.35$, $p=0.002$ between CON and SCHZ for global metastability.

#### Local Metastability

Global metastability reflects the average metastability across the modes. We were interested to also assess the local metastability in the modes.

For the HCPEP dataset we first investigated group (CON, NAP), run (RUN1, RUN2, RUN3, RUN4), and group x run effects on global META for each mode $\psi_1, \psi_2, \psi_3, \psi_4$, $\psi_5$). Using a 2x4 non-parametric ANOVA with the Aligned Rank Transform (ART)

[53], [54], we found significant interactions for run x group in $\psi_4$ $F(3,399)=3.665$, $p=0.013$, $\psi_5$ $F(3,399)=11.325$, $p<0.001$.

#### Main effects of run

In the CON group, we found significant main effects of run in $\psi_5$, $\chi^2 = 9$, $p<0.03$. In the NAP group, we found significant main effects of run in $\psi_4$, $\chi^2 = 19.77$, $p<0.001$ and in $\psi_5$, for $\chi^2 = 21.81$, $p<0.001$. The drivers for these effects and the associated effect sizes are detailed in S2 Supplementary Information.

#### Main effects of group

We found significant main effects of group in modes $\psi_4$ and $\psi_5$. The effect sizes of these differences were compared to the largest effect size between any pair of runs for that mode ($\psi_4$, *effect size*=0.308, $\psi_5$, *effect size*=0.410). We retained only group differences that were greater these run effects. We thus found significant group differences for $\psi_5$ in RUN1 ($p<0.001$, *effect size*=0.442).

We found a significant main effect of group for $\psi_1$, $p=0.0361$, *effect size*=0.090. There were no significant main effects of group for $\psi_2$ and $\psi_3$.

We thus inferred that mode metastability differed between CON and NAP in $\psi_1$ in all runs, and in $\psi_5$ in RUN1.

For the Cobre dataset, we found significant differences in mode META between CON and SCHZ in $\psi_3$ $t(128)=-4.69$, $p<0.001$, and $\psi_5$ $t(122)=-3.550$, $p=0.003$.

Complete statistical results for each dataset can be found in S2 Supporting Information. Figure 3 provides an overview of mode metastability data for each dataset in the form of raincloud plots.

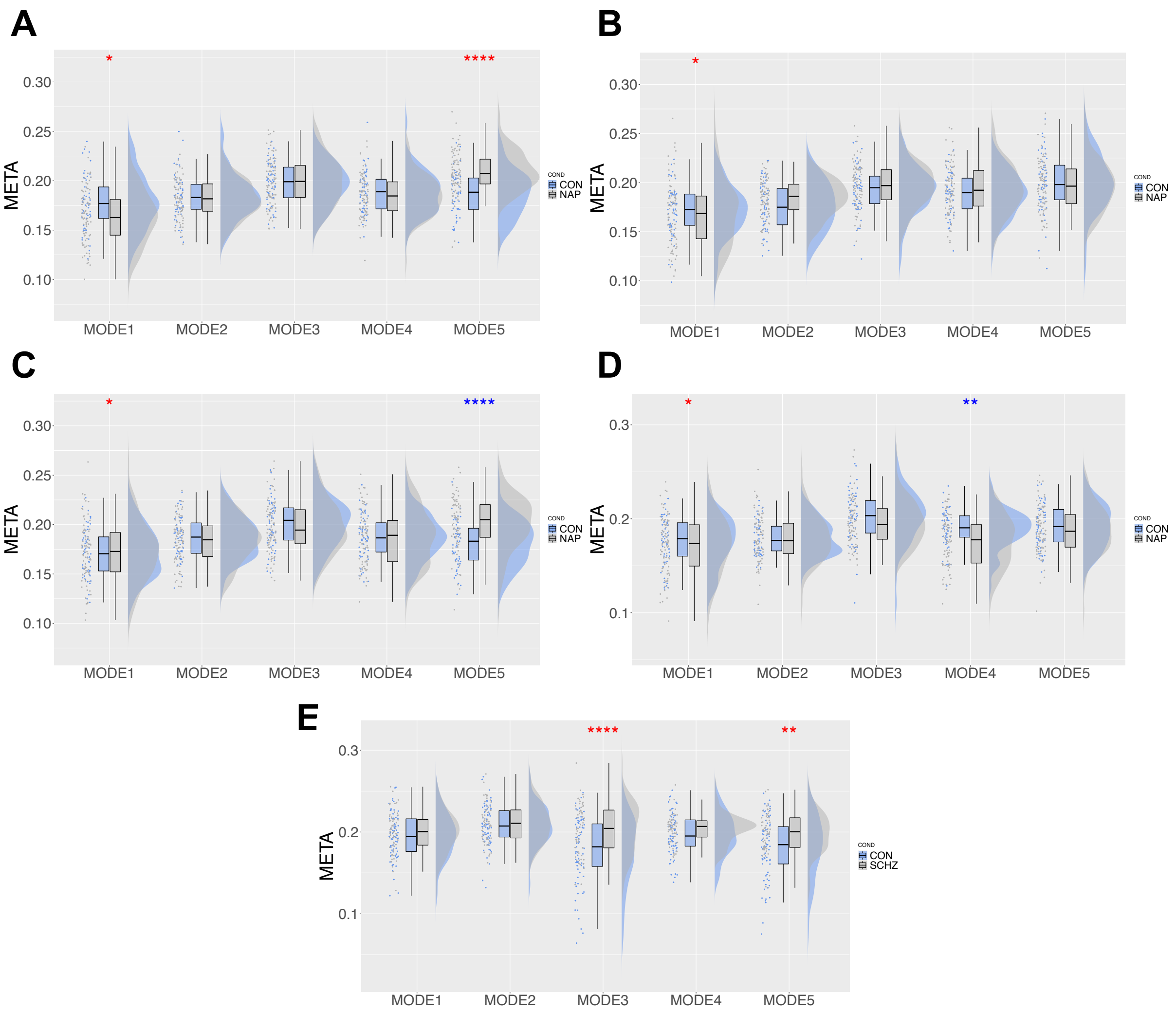


**Fig 3. Raincloud plots for META in each mode for HCPEP and Cobre datasets.** Raincloud plots show from left to right the raw data, boxplots showing the median, upper and lower quartiles, upper and lower extremes, and the distributions of the raw data. **A)** HCPEP RUN1. **B)** HCPEP RUN2. **C)** HCPEP RUN3. **D)** HCPCP RUN4. **E)** Cobre dataset. *=0.05, **=0.01,***=0.001,****<0.001. Red * effect size between groups greater than effect size between runs. Blue * effect size between groups less than largest effect size between runs.

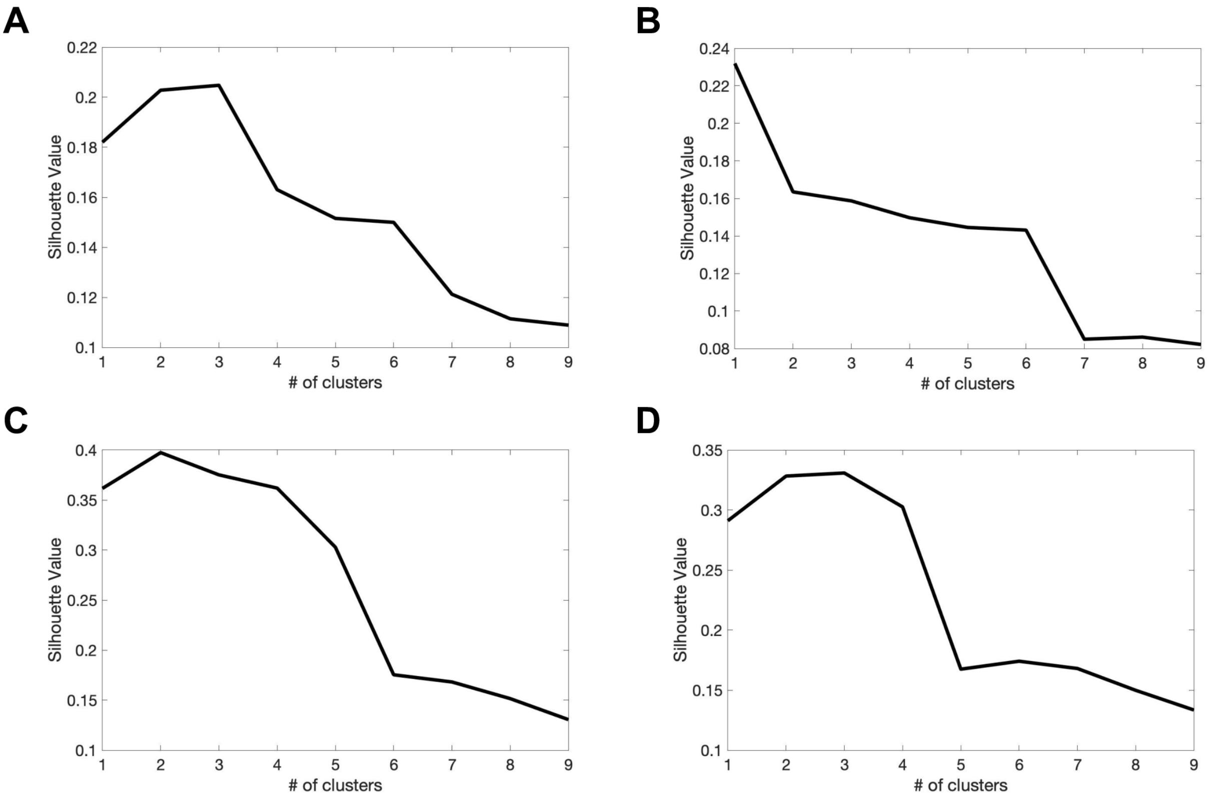


**S1 Fig Silhouette values for clustering solutions for 1 to 9 clusters with 2-10 modes respectively.** (**A**) HCPEP CON. (**B**) HCPEP NAP. (**C**) Cobre CON **(D)** Cobre SCHZ.

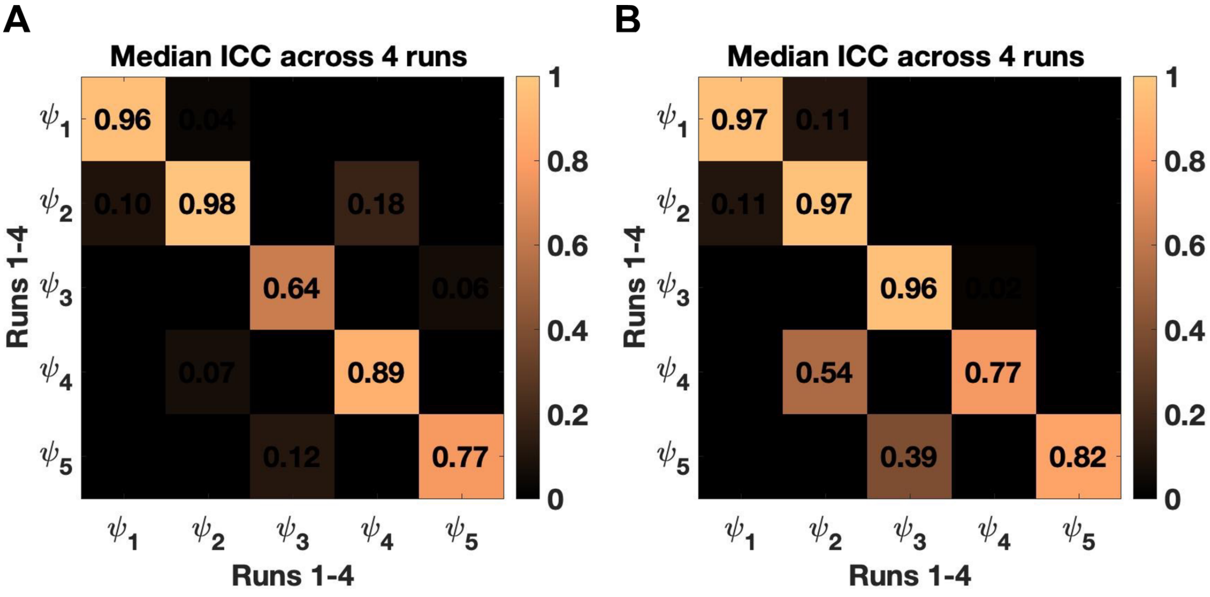


**S2 Fig Reliability of mode extraction for controls and non-affective psychosis**

### Bedrosian – Carson's empirical rule 'useful bandwidth $B_{\varphi}$'

$$B_{\varphi} = 2(\text{Modulation Index} + 1)f_m$$

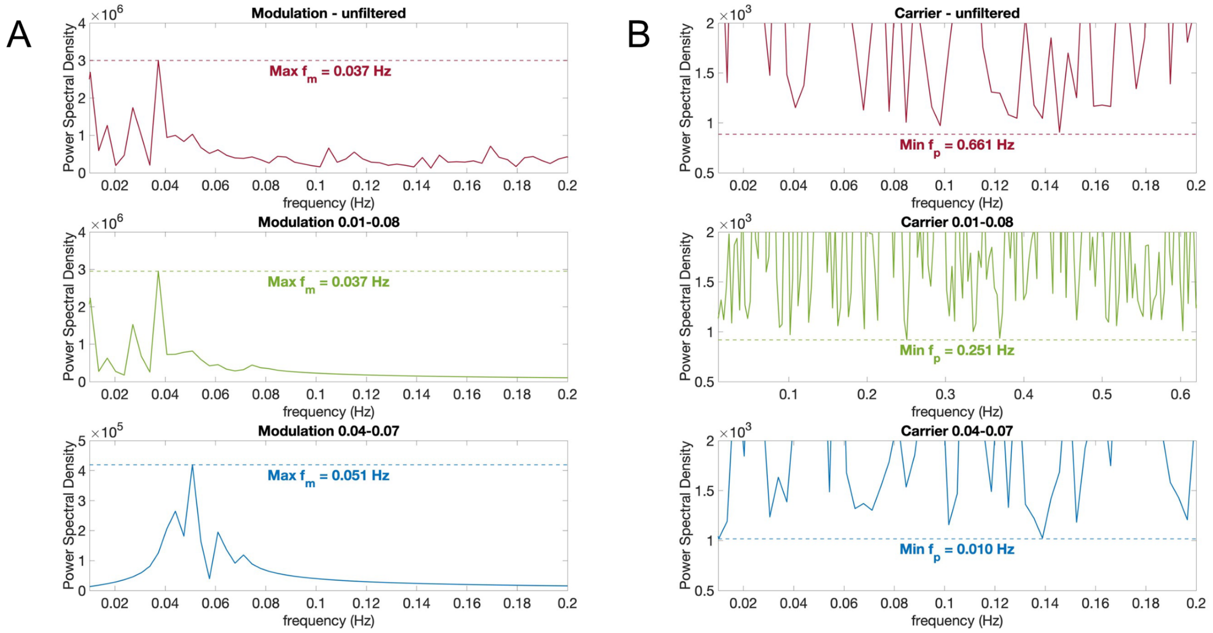


C

| | Max $f_m$ - $Freq_{Amplitude}$ | Modulation Index | Useful bandwidth $B_{\varphi}$ | Min $f_p$ - $Freq_{Phase}$ |
|---|---|---|---|---|
| unfiltered | 0.037 Hz | 0.590 | 0.119 Hz | 0.661 Hz |
| 0.01-0.08 | 0.037 Hz | 0.612 | 0.120 Hz | 0.251 Hz |
| 0.04-0.07 | 0.051 Hz | 0.585 | 0.161 Hz | 0.010 Hz |

**S3 Fig Non-violation of Bedrosian Theorem – HCPEP**

#### Bedrosian – Carson's empirical rule 'useful bandwidth $B_\varphi$'

$$B_\varphi = 2(\text{Modulation Index} + 1)f_m$$

A

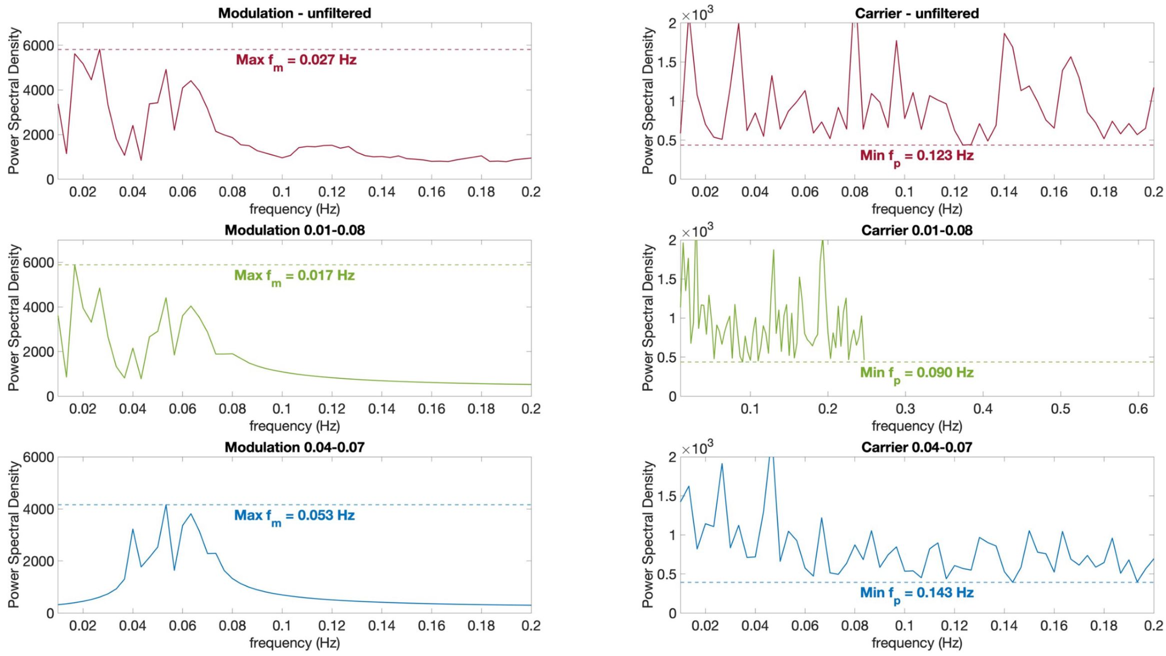


C

| | Max $f_m$ - $Freq_{Amplitude}$ | Modulation Index | Useful bandwidth $B_\varphi$ | Min $f_p$ - $Freq_{Phase}$ |
|---|---|---|---|---|
| unfiltered | 0.027 Hz | 0.616 | 0.086 Hz | 0.123 Hz |
| 0.01-0.08 | 0.017 Hz | 0.613 | 0.054 Hz | 0.090 Hz |
| 0.04-0.07 | 0.053 Hz | 0.613 | 0.172 Hz | 0.143 Hz |

**S4 Fig Non-violation of Bedrosian Theorem – Cobre**

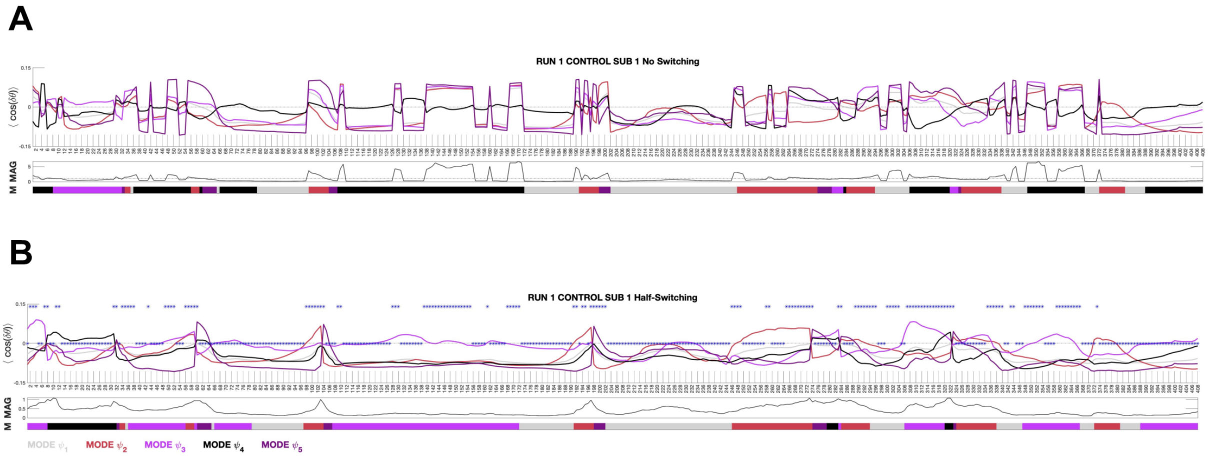


**S5 Fig Effect of smoothing on the leading eigenvector timeseries.** A) Timeseries for the leading eigenvectors for one subject without smoothing. B) Timeseries for the leading eigenvector for the same subject with half-switch smoothing. The blue asterixis indicate that half-switching occurred.

**S1 Table Assumption test results for contribution of basal ganglia regions FC in the HCPEP dataset.**

| | | Region | Mean | Stddev | Shapiro | p | Levine | p | df | Cohen's D | n1 | n2 | Magnitude |
|---|---|---|---|---|---|---|---|---|---|---|---|---|---|
| **HCPEP** | **CON** | **Caudate L** | 0.022 | 0.048 | 0.985 | 0.007 | 2.110 | 0.148 | (1,268) | 0.527 | 106 | 164 | moderate |
| | **NAP** | | -0.005 | 0.053 | | | | | | | | | |
| **HCPEP** | **CON** | **Caudate R** | 0.029 | 0.047 | 0.981 | 0.001 | 3.550 | 0.061 | (1,268) | 0.586 | 106 | 164 | moderate |
| | **NAP** | | -0.001 | 0.054 | | | | | | | | | |
| **HCPEP** | **CON** | **Putamen L** | 0.006 | 0.054 | 0.982 | 0.002 | 0.335 | 0.563 | (1,268) | 0.366 | 106 | 164 | small |
| | **NAP** | | -0.014 | 0.056 | | | | | | | | | |
| **HCPEP** | **CON** | **Putamen R** | -0.001 | 0.057 | 0.976 | 0.000 | 0.131 | 0.717 | (1,268) | 0.315 | 106 | 164 | small |
| | **NAP** | | -0.019 | 0.058 | | | | | | | | | |
| **HCPEP** | **CON** | **Pallidum L** | 0.003 | 0.048 | 0.994 | 0.305 | 0.006 | 0.938 | (1,268) | 0.309 | 106 | 164 | small |
| | **NAP** | | -0.012 | 0.048 | | | | | | | | | |
| **HCPEP** | **CON** | **Pallidum R** | -0.001 | 0.049 | 0.988 | 0.024 | 0.012 | 0.913 | (1,268) | 0.103 | 106 | 164 | negligible |
| | **NAP** | | -0.006 | 0.047 | | | | | | | | | |
| **HCPEP** | **CON** | **Thalamus L** | -0.009 | 0.055 | 0.962 | 0.000 | 5.559 | 0.019 | (1,268) | 0.647 | 106 | 164 | moderate |
| | **NAP** | | -0.041 | 0.048 | | | | | | | | | |
| **HCPEP** | **CON** | **Thalamus R** | -0.019 | 0.055 | 0.961 | 0.000 | 2.052 | 0.153 | (1,268) | 0.388 | 106 | 164 | small |
| | **NAP** | | -0.039 | 0.052 | | | | | | | | | |

We assessed the normality of the distribution of contribution with a Shapiro-Wilk test, equivalence of variance with a Levine test, and effect size with Cohen's D test.

**S2 Table Assumption test results for global META in the HCPEP and Cobre datasets.**

**Assumption Checks**

| | | Mean | Stddev | Shapiro | p | Levine | p | df | Cohen's D | n1 | n2 | Magnitude |
|---|---|---|---|---|---|---|---|---|---|---|---|---|
| **HCPEP META** | **CON** | 0.186 | 0.014 | 0.997 | 0.402 | 5.567 | 0.019 | (1,538) | -0.011 | 212 | 328 | negligible |
| | **NAP** | 0.186 | 0.016 | | | | | | | | | |

| | | Mean | Stddev | Shapiro | p | Levine | p | df | Cohen's D | n1 | n2 | Magnitude |
|---|---|---|---|---|---|---|---|---|---|---|---|---|
| **COBRE META** | **CON** | 0.192 | 0.023 | 0.942 | 0.000 | 2.673 | 0.105 | (1,128) | -0.573 | 71 | 59 | moderate |
| | **SCHZ** | 0.204 | 0.017 | | | | | | | | | |

We assessed the normality of the distribution of META with a Shapiro-Wilk test, equivalence of variance with a Levine test, and effect size with Cohen's D test.

**S3 Table. Assumption test results for metastability in the modes in the HCPEP and Cobre datasets.**

| Assumption Checks | | | | | | | | | | | | | |
|---|---|---|---|---|---|---|---|---|---|---|---|---|---|
| | | **Mode** | **Mean** | **Stddev** | **Shapiro** | **p** | **Levine** | **p** | **df** | **Cohen's D** | **n1** | **n2** | **Magnitude** |
| **HCPEP** | **CON** | **1** | 0.174 | 0.026 | 0.998 | 0.599 | 6.205 | 0.013 | (1,538) | 0.179 | 212 | 328 | negligable |
| **META** | **NAP** | | 0.169 | 0.030 | | | | | | | | | |
| **HCPEP** | **CON** | **2** | 0.181 | 0.022 | 0.998 | 0.881 | 0.276 | 0.600 | (1,538) | -0.042 | 212 | 328 | negligable |
| **META** | **NAP** | | 0.182 | 0.021 | | | | | | | | | |
| **HCPEP** | **CON** | **3** | 0.198 | 0.024 | 0.998 | 0.708 | 0.243 | 0.623 | (1,538) | 0.032 | 212 | 328 | negligable |
| **META** | **NAP** | | 0.197 | 0.024 | | | | | | | | | |
| **HCPEP** | **CON** | **4** | 0.188 | 0.021 | 0.998 | 0.888 | 10.422 | 0.001 | (1,538) | 0.117 | 212 | 328 | negligable |
| **META** | **NAP** | | 0.185 | 0.026 | | | | | | | | | |
| **HCPEP** | **CON** | **5** | 0.190 | 0.025 | 0.997 | 0.466 | 0.037 | 0.848 | (1,538) | -0.333 | 212 | 328 | small |
| **META** | **NAP** | | 0.198 | 0.026 | | | | | | | | | |
| | | **Mode** | **Mean** | **Stddev** | **Shapiro** | **p** | **Levine** | **p** | **df** | **Cohen's D** | **n1** | **n2** | **Magnitude** |
| **COBRE** | **CON** | **1** | 0.195 | 0.027 | 0.990 | 0.442 | 0.608 | 0.437 | (1,128) | -0.175 | 71 | 59 | negligible |
| **META** | **SCHZ** | | 0.200 | 0.025 | | | | | | | | | |
| **COBRE** | **CON** | **2** | 0.208 | 0.025 | 0.991 | 0.601 | 0.006 | 0.940 | (1,128) | -0.133 | 71 | 59 | negligible |
| **META** | **SCHZ** | | 0.211 | 0.024 | | | | | | | | | |
| **COBRE** | **CON** | **3** | 0.177 | 0.041 | 0.963 | 0.001 | 3.648 | 0.058 | (1,128) | -0.716 | 71 | 59 | moderate |
| **META** | **SCHZ** | | 0.204 | 0.031 | | | | | | | | | |
| **COBRE** | **CON** | **4** | 0.197 | 0.025 | 0.989 | 0.412 | 9.397 | 0.003 | (1,128) | -0.319 | 71 | 59 | small |
| **META** | **SCHZ** | | 0.204 | 0.018 | | | | | | | | | |
| **COBRE** | **CON** | **5** | 0.180 | 0.038 | 0.957 | 0.000 | 9.246 | 0.003 | (1,128) | -0.603 | 71 | 59 | moderate |
| **META** | **SCHZ** | | 0.200 | 0.025 | | | | | | | | | |

We assessed the normality of the distribution of mode META with a Shapiro-Wilk test, equivalence of variance with a Levine test, and effect size with Cohen's D test.

**S4 Table. Assumption test results for global VAR in the HCPEP and Cobre datasets**.

| Assumption Checks | | | | | | | | | | | | |
|---|---|---|---|---|---|---|---|---|---|---|---|---|
| | | **Mean** | **Stddev** | **Shapiro** | **p** | **Levine** | **p** | **df** | **Cohen's D** | **n1** | **n2** | **Magnitude** |
| **HCPEP** | **CON** | 0.006 | 0.001 | 0.960 | 0.000 | 0.124 | 0.725 | (1,538) | -0.246 | 212 | 328 | small |
| **VAR** | **NAP** | 0.006 | 0.001 | | | | | | | | | |
| | | **Mean** | **Stddev** | **Shapiro** | **p** | **Levine** | **p** | **df** | **Cohen's D** | **n1** | **n2** | **Magnitude** |
| **COBRE** | **CON** | 0.004 | 0.001 | 0.985 | 0.148 | 0.121 | 0.728 | (1,128) | -0.730 | 71 | 59 | moderate |
| **VAR** | **SCHZ** | 0.005 | 0.001 | | | | | | | | | |

We assessed the normality of the distribution of META with a Shapiro-Wilk test, equivalence of variance with a Levine test, and effect size with Cohen's D test.

**S5 Table 7. Assumption test results for mode VAR in the HCPEP and Cobre datasets**.

**Assumption Checks**

| | | Mode | Mean | Stddev | Shapiro | p | Levine | p | df | Cohen's D | n1 | n2 | Magnitude |
|---|---|---|---|---|---|---|---|---|---|---|---|---|---|
| **HCPEP** | **CON** | **1** | 0.0058 | 0.0012 | 0.958 | 0.000 | 0.179 | 0.673 | (1,538) | -0.220 | 212 | 328 | small |
| **VAR** | **NAP** | | 0.0061 | 0.0012 | | | | | | | | | |
| **HCPEP** | **CON** | **2** | 0.0065 | 0.0012 | 0.954 | 0.000 | 0.027 | 0.870 | (1,538) | -0.206 | 212 | 328 | small |
| **VAR** | **NAP** | | 0.0068 | 0.0012 | | | | | | | | | |
| **HCPEP** | **CON** | **3** | 0.0057 | 0.0014 | 0.965 | 0.000 | 0.785 | 0.376 | (1,538) | -0.106 | 212 | 328 | negligible |
| **VAR** | **NAP** | | 0.0059 | 0.0015 | | | | | | | | | |
| **HCPEP** | **CON** | **4** | 0.0060 | 0.0011 | 0.978 | 0.000 | 1.677 | 0.196 | (1,538) | -0.363 | 212 | 328 | small |
| **VAR** | **NAP** | | 0.0064 | 0.0012 | | | | | | | | | |
| **HCPEP** | **CON** | **5** | 0.0055 | 0.0015 | 0.983 | 0.000 | 2.426 | 0.120 | (1,538) | -0.281 | 212 | 328 | small |
| **VAR** | **NAP** | | 0.0059 | 0.0014 | | | | | | | | | |
| | | **Mode** | **Mean** | **Stddev** | **Shapiro** | **p** | **Levine** | **p** | **df** | **Cohen's D** | **n1** | **n2** | **Magnitude** |
| **COBRE** | **CON** | **1** | 0.0037 | 0.0013 | 0.984 | 0.144 | 0.000 | 0.983 | (1,128) | -0.601 | 71 | 59 | moderate |
| **VAR** | **SCHZ** | | 0.0044 | 0.0012 | | | | | | | | | |
| **COBRE** | **CON** | **2** | 0.0045 | 0.0014 | 0.990 | 0.468 | 0.932 | 0.336 | (1,128) | -0.576 | 71 | 59 | moderate |
| **VAR** | **SCHZ** | | 0.0053 | 0.0012 | | | | | | | | | |
| **COBRE** | **CON** | **3** | 0.0034 | 0.0016 | 0.975 | 0.018 | 0.000 | 0.994 | (1,128) | -0.631 | 71 | 59 | moderate |
| **VAR** | **SCHZ** | | 0.0044 | 0.0016 | | | | | | | | | |
| **COBRE** | **CON** | **4** | 0.0040 | 0.0013 | 0.991 | 0.616 | 0.013 | 0.908 | (1,128) | -0.756 | 71 | 59 | moderate |
| **VAR** | **SCHZ** | | 0.0049 | 0.0012 | | | | | | | | | |
| **COBRE** | **CON** | **5** | 0.0031 | 0.0015 | 0.989 | 0.362 | 2.503 | 0.116 | (1,128) | -0.821 | 71 | 59 | large |
| **VAR** | **SCHZ** | | 0.0042 | 0.0012 | | | | | | | | | |

We assessed the normality of the distribution of VAR in each mode with a Shapiro-Wilk test, equivalence of variance with a Levine test, and effect size with Cohen's D test.

**S6 Table. Neurosynth terms**

| | | | |
|---|---|---|---|
| action | episodic_memory | memory_retrieval | semantic_knowledge |
| adaptation | expectancy | mental_imagery | semantic_memory |
| addiction | expertise | monitoring | sentence_comprehension |
| anticipation | face_recognition | mood | skill |
| anxiety | facial_expression | morphology | sleep |
| arousal | familiarity | motor_control | social_cognition |
| association | fear | movement | spatial_attention |
| attention | fixation | multisensory | speech_perception |
| attentional_resources | focus | naming | speech_production |
| autobiographical_memory | gaze | navigation | strategy |
| balance | goal | object_recognition | strength |
| belief | hyperactivity | pain | stress |
| categorization | imagery | perception | sustained_attention |
| cognitive_control | impulsivity | planning | task_difficulty |
| communication | induction | priming | task_switching |
| competition | inference | psychosis | thought |
| concept | inhibition | reading | timing |
| consciousness | insight | reasoning | uncertainty |
| consolidation | integration | recall | updating |
| context | intelligence | recognition | utility |
| coordination | intention | rehearsal | valence |
| cueing | interference | response_conflict | verbal_fluency |
| decision | judgment | response_inhibition | verbal_memory |
| decision_making | knowledge | response_selection | visual_attention |
| detection | language | retention | visual_perception |
| discrimination | language_comprehension | retrieval | visual_search |
| distraction | learning | reward_anticipation | word_recognition |
| eating | listening | rhythm | working_memory |
| efficiency | localization | risk | |
| effort | loss | rule | |
| emotion | maintenance | salience | |
| emotion_regulation | manipulation | search | |
| empathy | meaning | selective_attention | |
| encoding | memory | semantic_information | |

**S1 Supplementary Information**

| **Region of Interest** | **Main effect of group** | | | **Main effect of run (largest)** | | | **Interaction Group x Run** | |
|---|---|---|---|---|---|---|---|---|
| | **Z** | **p** | **effect size** | **Z** | **p** | **effect size** | **F** | **p** |
| Caudate_L | 3296 | **<0.001** | 0.435 | 1179 | **<0.001** | 0.400 | F(3,339) = 4.899 | **0.002** |
| Caudate_R | 3521 | **<0.001** | 0.523 | 656 | **<0.001** | 0.382 | F(3,339) = 7.718 | **<0.001** |
| Putamen_L | 3079 | **<0.001** | 0.351 | 661 | **<0.001** | 0.335 | F(3,339) = 7.923 | **<0.001** |
| Putamen_R | 3093 | **<0.001** | 0.357 | 668 | **<0.001** | 0.333 | F(3,339) = 5.297 | **0.001** |
| Pallidum_L | 2804 | **0.005** | 0.245 | 1008 | **0.008** | 0.266 | F(3,339) = 3.394 | **0.018** |
| Pallidum_R | 38175 | 0.054 | 0.083 | | | | F(3,339) = 1.673 | 0.083 |
| Thalamus_L | 3530 | **<0.001** | 0.526 | 1132 | **0.001** | 0.360 | F(3,339) = 8.105 | **<0.001** |
| Thalamus_R | 3038 | **<0.001** | 0.335 | 635 | **<0.001** | 0.349 | F(3,339) = 3.032 | **0.029** |

Results from statistical tests for differences in basal ganglia connectivity between HC and NAP in the HCPEP dataset

**S2 Supplementary Information**

**META**

**Step 1** **2x4 non-parametric ANOVA using ART**

| | Term | F | Df | Df.res | Pr(>F) |
|---|---|---|---|---|---|
| COND | COND | 0.235 | 1 | 133 | 0.629 |
| RUN | RUN | 1.302 | 3 | 399 | 0.273 |
| COND:RUN | COND:RUN | 3.192 | 3 | 399 | 0.024 |

**Step 2** **CON:1x4 repeated measures non-parametric ANOVA using Friedmann test**

| chi-squared | df | p.value | method | data |
|---|---|---|---|---|
| 0.872 | 3 | 0.832 | Friedman | META and RUN and SUB |

**Step 4** **NAP:1x4 repeated measures non-parametric ANOVA using Friedmann test**

| chi-squared | df | p.value | method | data |
|---|---|---|---|---|
| 6.673 | 3 | 0.083 | Friedman | META and RUN and SUB |

**Step 6** **Non-parametric Wilcox test between CON and NAP for each run**

| | .y. | group1 | group2 | n1 | n2 | statistic | p | p.signif | effsize | magnitude |
|---|---|---|---|---|---|---|---|---|---|---|
| RUN1 | META | CON | NAP | 53 | 82 | 2082 | 0.683 | ns | 0.035 | small |
| RUN2 | META | CON | NAP | 53 | 82 | 2131 | 0.852 | ns | 0.016 | small |
| RUN3 | META | CON | NAP | 53 | 82 | 2003 | 0.445 | ns | 0.066 | small |
| RUN4 | META | CON | NAP | 53 | 82 | 2694 | 0.019 | * | 0.202 | small |

Results from statistical tests for differences in mode META between HC and NAP in the HCPEP dataset, and HC and SCHZ in the Cobre dataset

#### S3 Supplementary Information

**VAR**

**Step 1** **2x4 non-parametric ANOVA using ART**

| | Term | F | Df | Df.res | Pr(>F) |
|---|---|---|---|---|---|
| COND | COND | 4.251 | 1 | 133 | 0.041 |
| RUN | RUN | 1.499 | 3 | 399 | 0.214 |
| COND:RUN | COND:RUN | 8.411 | 3 | 399 | 0.000 |

**Step 2** **CON:1x4 repeated measures non-parametric ANOVA using Friedmann test**

| chi-squared | df | p.value | method | data |
|---|---|---|---|---|
| 3.136 | 3 | 0.371 | Friedman | VAR and RUN and SUB |

**Step 4** **NAP:1x4 repeated measures non-parametric ANOVA using Friedmann test**

| chi-squared | df | p.value | method | data |
|---|---|---|---|---|
| 19.156 | 3 | 0.000 | Friedman | VAR and RUN and SUB |

**Step 5** **NAP:Non-parametric pair-wise comparisons using paired Wilcox test**

| | group1 | group2 | n1 | n2 | statistic | p | p.adj | p.a.sig | effsize | magnitude |
|---|---|---|---|---|---|---|---|---|---|---|
| VAR | RUN1 | RUN2 | 82 | 82 | 1798 | 0.657 | 1.000 | ns | 0.051 | small |
| VAR | RUN1 | RUN3 | 82 | 82 | 2413 | 0.001 | 0.006 | ** | 0.215 | small |
| VAR | RUN1 | RUN4 | 82 | 82 | 2492 | 0.000 | 2492 | ** | 0.216 | small |
| VAR | RUN2 | RUN3 | 82 | 82 | 2244 | 0.012 | 0.073 | ns | 0.157 | small |
| VAR | RUN2 | RUN4 | 82 | 82 | 2249 | 0.011 | 0.068 | ns | 0.165 | small |
| VAR | RUN3 | RUN4 | 82 | 82 | 1910 | 0.336 | 1.000 | ns | 0.029 | small |

**Step 6** **Non-parametric Wilcox test between CON and NAP for each run**

| | group1 | group2 | n1 | n2 | statistic | p | p.signif | effsize | magnitude |
|---|---|---|---|---|---|---|---|---|---|
| RUN1 | CON | NAP | 53 | 82 | 1455 | 0.001 | ** | 0.278 | small |
| RUN2 | CON | NAP | 53 | 82 | 1494 | 0.002 | ** | 0.263 | small |
| RUN3 | CON | NAP | 53 | 82 | 2140 | 0.884 | ns | 0.013 | small |
| RUN4 | CON | NAP | 53 | 82 | 2326 | 0.492 | ns | 0.059 | small |

Results from statistical tests for differences in mode VAR between HC and NAP in the HCPEP dataset, and HC and SCHZ in the Cobre dataset
